# Utility of skin tone on pulse oximetry in critically ill patients: a prospective cohort study

**DOI:** 10.1101/2024.02.24.24303291

**Authors:** Sicheng Hao, Katelyn Dempsey, João Matos, Christopher E. Cox, Veronica Rotemberg, Judy W. Gichoya, Warren Kibbe, Chuan Hong, Ian Wong

**Author notes:** co-first authors (Sicheng Hao, Katelyn Dempsey). **Corresponding author:** A. Ian Wong, MD, PhD, Assistant Professor, Department of Medicine, Division of Pulmonary, Allergy, and Critical Care Medicine, Department of Biostatistics and Bioinformatics, Division of Translational Biomedical Informatics, Duke University, 2 Genome Court, Box 103000, Durham, NC 27710.

## Abstract

**Importance:** Pulse oximetry, a ubiquitous vital sign in modern medicine, has inequitable accuracy that disproportionately affects Black and Hispanic patients, with associated increases in mortality, organ dysfunction, and oxygen therapy. Although the root cause of these clinical performance discrepancies is believed to be skin tone, previous retrospective studies used self-reported race or ethnicity as a surrogate for skin tone.

**Objective:** To determine the utility of objectively measured skin tone in explaining pulse oximetry discrepancies.

**Design, Setting, and Participants:** Admitted hospital patients at Duke University Hospital were eligible for this prospective cohort study if they had pulse oximetry recorded up to 5 minutes prior to arterial blood gas (ABG) measurements. Skin tone was measured across sixteen body locations using administered visual scales (Fitzpatrick Skin Type, Monk Skin Tone, and Von Luschan), reflectance colorimetry (Delfin SkinColorCatch [L*, individual typology angle {ITA}, Melanin Index {MI}]), and reflectance spectrophotometry (Konica Minolta CM-700D [L*], Variable Spectro 1 [L*]).

**Main Outcomes and Measures:** Mean directional bias, variability of bias, and accuracy root mean square (A_RMS_), comparing pulse oximetry and ABG measurements. Linear mixed-effects models were fitted to estimate mean directional bias while accounting for clinical confounders.

**Results:** 128 patients (57 Black, 56 White) with 521 ABG–pulse oximetry pairs were recruited, none with hidden hypoxemia. Skin tone data was prospectively collected using 6 measurement methods, generating 8 measurements. The collected skin tone measurements were shown to yield differences among each other and overlap with self-reported racial groups, suggesting that skin tone could potentially provide information beyond self-reported race. Among the eight skin tone measurements in this study, and compared to self-reported race, the Monk Scale had the best relationship with differences in pulse oximetry bias (point estimate: −2.40%; 95% CI: −4.32%, - 0.48%; *p=*0.01) when comparing patients with lighter and dark skin tones.

**Conclusions and relevance:** We found clinical performance differences in pulse oximetry, especially in darker skin tones. Additional studies are needed to determine the relative contributions of skin tone measures and other potential factors on pulse oximetry discrepancies.

**Key Points:** *Question:* Can skin tone capture information beyond race to help explain pulse oximetry discrepancies?

*Findings:* Pulse oximetry bias across races seems to persist across skin tone when measured using administered visual scales, reflectance colorimetry, or reflectance spectrophotometry. Among the eight skin tone measurements in this study, and compared to self-reported race, the Monk Scale seemed to best correlate with pulse oximetry bias when comparing patients with lighter and dark skin tones.

*Meaning:* Compared to self-reported race, skin tone is associated with some pulse oximetry discrepancies; we recommend using skin tone to assist the regulatory clearance of equitable pulse oximeters.

## Background

Racial and ethnic bias in pulse oximetry stands out as a quintessential health inequity, whereby the same medical devices that guide clinical decision-making may fail to function equally well for all patients.^1^ The reliability of pulse oximetry has been a reason for concern for decades,^2–6^ but it was not until the COVID-19 pandemic when Sjoding and colleagues’ seminal paper reported racial bias in pulse oximetry measurements that pulse oximetry became a health equity issue.^7^

Followed by other studies,^8–15^ oxygen saturation measured by pulse oximetry (SpO_2_) is widely reported to overestimate the “true” arterial oxygen (SaO_2_), measured by arterial blood gas (ABG), disproportionately affecting Black and Hispanic patients. A seemingly small discrepancy is associated with higher rates of “hidden hypoxemia” among these patients,^2, 8, 12^ with associated inequities in oxygen therapies ^9, 16^ and increases in mortality and organ dysfunction.^8^

Pulse oximeters estimate arterial oxygen saturation by measuring light absorption of oxyhemoglobin and deoxyhemoglobin in capillary blood.^17, 18^ Previous studies have shown that skin tone can independently affect light absorption, causing discrepant readings, especially among darker-skinned individuals. ^19–21^As such, previous retrospective studies share a fundamental limitation: self-reported race or ethnicity is used as a surrogate for skin tone, although the root cause of these discrepancies is believed to be skin tone.^22^

In this cohort study, we prospectively collected skin tone data from critically ill patients in various body locations using different devices. We paired this data with pulse oximetry measurements, ABG, and other Electronic Health Records (EHR) data to investigate the utility of skin tone data in explaining pulse oximetry performance.

As a pilot study, our objectives were 2-fold: first, to provide a framework to conduct larger clinical studies that assess the association between different skin tone measurements and pulse oximetry discrepancies; and second, to provide evidence that can support recent discussions from the Food and Drug Administration (FDA)^23–26^ in pursuit of guidelines to evaluate pulse oximetry performance in a more inclusive spectrum of patients.

Pulse oximetry performance is commonly assessed by the FDA using the accuracy root mean square (A_RMS_), with a threshold of A_RMS_ ≤ 3.0% for transmittance pulse oximeters between SaO_2_ and SpO_2_ measurements ranging from 70–100%.^27^ A_RMS_ can be decomposed into the average difference in magnitude between SaO_2_ and SpO_2,_ often referred to as “bias”, and the variability of those differences, which can be measured as the standard deviation of the difference between SaO_2_ and SpO_2_.

## Methods

This study was approved by the Duke Health IRB under Pro00110842, following the American Medical Association’s recommendations on health equity language and adhering to the STROBE statement.^28, 29^

### Cohort Selection

Patients admitted to an adult intensive care unit (ICU) at Duke University Hospital were screened. Standard-of-care ABG up to 5 minutes after a pulse oximetry measurement was required for eligibility, resulting in SaO_2_–SpO_2_ pairs.

Exclusion criteria included unremovable fingernail polish, admission for a vascular complication (e.g., grafting or stenting), amputation, and large areas of skin discoloration where the accuracy of skin tone measurements could be affected. Pairs containing either a SaO_2_ or a SpO_2_ measurement out of the 70–100% range were excluded.^8^ ^27^

### Data Collection and Processing

All data and patients’ consent were stored in Duke Health’s REDCap, with data processing performed in Python 3.10.

The mathematical definitions of mean directional bias, variability of bias, and A_RMS_ are in Supplemental Formulas 2, 3, and 4.

### Skin Tone

Three types of skin tone assessment were conducted using different devices: administered visual scales (Fitzpatrick, Monk^30^, and Von Luschan scales, visible in Supplemental Figures 6a, 6b and 6c); reflectance colorimetry (Delfin SkinColorCatch); and reflectance spectrophotometry (Variable Spectro 1 Pro Bridge Set and Konica Minolta CM-700D Spectrophotometer).

All the skin tone data are collected within 7 days of the SaO_2_–SpO_2_ pairs and with controlled lighting to ensure reproducibility. Using the L*, individual typology angle (ITA) and Melanin Index (Melanin Index) color spaces, eight different skin tone measures were collected in this study, as detailed in Supplemental Table 2. Further details are demonstrated in Supplemental Text.

### Data merging

Pulse oximetry values and ABG panel data were merged into SaO_2_-SpO_2_ pairs and recorded in REDCap. Demographic data was merged from the EHR system. Three race groups were defined, including “Black”, “Other” and “White” patients. The group “Other” captures minority patients who self-identify as Asian, American Indian / Alaskan natives, more than two races, and unknown race–groups that separately represent a small proportion of patients. Vital signs captured within 4 hours prior to the SaO_2_–SpO_2_ pair were merged from the EHR system. Mean arterial pressure (MAP) from the arterial line was preferred when available, otherwise cuff values were used. Laboratory test values from the previous 24 hours, relative to the SaO_2_– SpO_2_ pair, were merged (listed in Supplemental Table 1).

### Missingness

Missing data occurred occasionally in two skin tone measurements, Variable L* and Konica Minolta L*, due to technical issues or patient refusal. Missingness rates for skin tone measurements can be found in Table 1. In the merged EHR clinical data, missingness occurred in vital signs and laboratory test values when no value was found within the set windows. Missingness rates for these covariates are in Supplemental Table 1. Patients with missing data that would be necessary for modeling were dropped from the study, and sensitivity analyses were run to assess the robustness of this design choice.

**Table 1.** Characteristics of the cohort obtained after applying inclusion and exclusion criteria, grouped by race. Demographic information for all 128 patients, along with their skin tone measurements, were grouped by race. The group “Other” contains patients who self-identify as Asian (n= 5), American Indian / Alaskan natives (n= 6), More than two races (n= 2), and Unknown race (n= 2). Among the eight skin tone scales, Monk scale, Fitzpatrick scale, Von Luschan scale, and Delfin Melanin Index are ordered numerically ascending from light to dark, the other ones are ascending.

|  |  | Patients grouped by race group |  |  |  |  |
| --- | --- | --- | --- | --- | --- | --- |
|  |  | Missing | Black | Other | White | Overall |
| **n** |  |  | 57 | 15 | 56 | 128 |
| **Ethnicity, n (%)** | **Not Hispanic/Latino** | 0 | 57 (100.0) | 12 (80.0) | 52 (92.9) | 121 (94.5) |
|  | **Hispanic/Latino** |  |  | 1 (6.7) | 3 (5.4) | 4 (3.1) |
|  | **Unknown** |  |  | 2 (13.3) | 1 (1.8) | 3 (2.3) |
| **Gender, n (%)** | **Female** | 0 | 23 (40.4) | 3 (20.0) | 25 (44.6) | 51 (39.8) |
| **Observed oximeter location, n (%)** | **Forehead** | 0 | 1 (1.8) | 0 (0.0) | 0 (0.0) | 1 (0.8) |
|  | **Missing** |  | 18 (31.6) | 7 (46.7) | 20 (35.7) | 45 (35.2) |
|  | **Palm average** |  | 36 (63.2) | 7 (46.7) | 36 (64.3) | 79 (61.7) |
|  | **Right toe** |  | 2 (3.5) | 1 (6.7) | 0 (0.0) | 3 (2.3) |
| **Fitzpatrick scale, mean (SD)** |  | 0 | 4.8 (0.6) | 3.6 (1.1) | 2.7 (0.6) | 3.7 (1.2) |
| **Von Luschan scale, mean (SD)** |  | 0 | 27.3 (2.3) | 22.0 (5.2) | 19.0 (3.4) | 23.0 (5.1) |
| **Monk scale, mean (SD)** |  | 0 | 6.4 (0.6) | 5.2 (1.4) | 4.3 (0.7) | 5.3 (1.2) |
| **Delfin ITA, mean (SD)** |  | 0 | -2.9 (13.9) | 21.0 (12.5) | 37.2 (11.9) | 17.4 (22.8) |
| **Delfin L\*, mean (SD)** |  | 0 | 48.9 (4.8) | 56.4 (6.6) | 63.1 (4.3) | 56.0 (8.2) |
| **Variable L\*, mean (SD)** |  | 29 | 49.9 (5.1) | 57.7 (5.5) | 63.1 (4.0) | 57.1 (7.8) |
| **Konica Minolta L\*, mean (SD)** |  | 8 | 40.4 (4.9) | 48.2 (5.0) | 54.4 (4.6) | 47.5 (8.1) |
| **Delfin Melanin Index, mean (SD)** |  | 0 | 742.2 (42.3) | 650.2 (51.7) | 598.4 (49.4) | 668.5 (82.4) |

### Measurement variability

The standard deviation was computed across the different values of the same measure and location, and compared with the average standard deviations across all locations. Average standard deviations across palm and finger locations were also computed, as depicted in Supplemental Table 3.

### Statistical Analysis

Exploratory data analysis was performed using Python 3.10^31^, as described in the Supplemental Text, and summarized using the *tableone* package,^32^ in Table 1. For each tertile of skin tone, mean directional bias, variability of bias, and A_RMS_ were computed, as reported in Figure 2. Statistical analysis was conducted in R 4.3.1,^33^ using the packages *nlme* for the mixed-effects analysis.^34, 35^

### Linear mixed-effects models

Linear mixed-effects models with patient identifiers as a random effect were fitted and adjusted for potential confounders (race and clinical features). Pairs with missing data were dropped for the analysis.

As a baseline, we built a model to assess the effect of self-reported race in pulse oximetry mean directional bias, adjusting for pH, SaO_2_, heart rate (HR), and MAP (Supplemental Formula 5). The following models, documented in Supplemental Formula 6, included these same covariates, as well as the skin tone variables, separate per model (eight models were built in total to assess the individual effect of each skin tone variable).

Lastly, to investigate the combined effect of all the skin tone measurements on pulse oximeter mean directional bias, we fitted two linear mixed-effects models (Supplemental Formula 7 and Supplemental Formula 8). The first model included six skin tone variables, excluding Konica Minolta L* and Variable L* due to missingness. The second model included all eight skin tone measurements, as a sensitivity analysis. These differences in design resulted in a lower sample size for the second model. Table 2 summarizes the built models. All the significance levels in linear mixed-effects models are calculated using likelihood ratio tests (LRT) using chi-squared statistics.

**Table 2.** Results of the adjusted linear mixed-effects models. Results of the four linear mixed-effects models with clinical variables (SaO_2_, pH heart rate, and MAP) adjusted, (Supplemental Formulas 5 to 9). Likelihood ratio tests (LRT) are performed to demonstrate whether the null hypothesis should be rejected. Variables and coefficients are derived from the linear mixed-effects model with a negative value being a larger magnitude of bias, □2 statistics, and p-values are derived from LRT test results. N is the sample size of each model. Green cells represent negative coefficient values, i.e. the variable has an effect towards an overestimation of SaO_2_, and vice-versa for green cells. Bold, underlined p-values denote that the significance threshold was passed at 0.05 and the null hypothesis was rejected. Expected Total Effect^1^: The expected difference in estimated measurement bias of the darkest and lightest subject (assuming the normalized value of all skin tone measurements is 1 for the darkest subject and 0 for the lightest), computed as the sum of the separate coefficients in gray, above. The self-reported race alone (Supplemental Formula 5) presents coefficients in the expected direction (−0.23%, 95% CI: −0.76%, 0.30%; p-value: 0.64 for Black patients, compared to White patients), but the p-value is not significant. When assessing the effect of a separate skin tone scale on bias (Supplemental Formula 6), only the Monk skin tone scale is shown to be significant (−2.40%; 95% CI: −4.32%, −0.48%; p-value: 0.01). The effect of all combined six skin tone scales on bias (the ones without missingness, Supplemental Formula 7) was found to be significant, with an expected total effect ^1^ of −1.72%, p-value of 0.02. Finally, when considering all eight skin tone scale variables, this expected total effect remains in the expected direction (- 3.80%), although the p-value is not significant (p-value = 0.06).

| Model | Variables | Coefficients | Lower 95% CI | Upper 95% CI | Chi-Squared | P-value | N |
| --- | --- | --- | --- | --- | --- | --- | --- |
| Association between race and bias (Supplemental Formula 5) | White | Baseline |  |  | 0.90 | 0.64 | 463 |
|  | Black | -0.23 | -0.76 | 0.30 |  |  |  |
|  | Other | -0.31 | -1.31 | 0.69 |  |  |  |
| Association between separate skin tone scale and bias (Supplemental Formula 6) | Fitzpatrick | -0.75 | -2.07 | 0.57 | 1.37 | 0.24 | 463 |
|  | Von Luschan | -1.10 | -2.68 | 0.48 | 1.99 | 0.16 | 463 |
|  | Monk | -2.40 | -4.32 | -0.48 | 6.07 | **0.01** | 463 |
|  | Delfin ITA | -0.62 | -2.33 | 1.09 | 0.58 | 0.45 | 463 |
|  | Delfin L* | 0.06 | -1.70 | 1.82 | <0.001 | 0.98 | 463 |
|  | Konica Minolta L* | -0.77 | -2.58 | 1.04 | 0.76 | 0.38 | 424 |
|  | Variable L* | -0.31 | -2.03 | 1.41 | 0.18 | 0.67 | 367 |
|  | Delfin Melanin Index | 0.17 | -1.53 | 1.87 | 0.03 | 0.87 | 463 |
| Association between six of skin tone scales and bias (Supplemental Formula 7) | Fitzpatrick | 0.37 | -2.27 | 3.01 | 14.98 | **0.02** | 463 |
|  | Von Luschan | 0.36 | -2.53 | 3.25 |  |  |  |
|  | Monk | -3.55 | -6.93 | -0.17 |  |  |  |
|  | Delfin ITA | -6.62 | -12.22 | -1.02 |  |  |  |
|  | Delfin L* | 4.66 | -0.40 | 9.72 |  |  |  |
|  | Delfin Melanin Index | 3.06 | -1.15 | 7.27 |  |  |  |
|  | **Expected Total Effect<sup>1</sup>** | **-1.72** | -1.72 | -1.72 |  |  |  |
| Association between eight of skin tone scales and bias (Supplemental Formula 8) | Fitzpatrick | -0.76 | -4.00 | 2.48 | 14.86 | 0.06 | 328 |
|  | Von Luschan | 2.53 | -1.03 | 6.09 |  |  |  |
|  | Monk | -3.43 | -7.61 | 0.75 |  |  |  |
|  | Delfin ITA | -7.61 | -14.40 | -0.82 |  |  |  |
|  | Delfin L* | 3.58 | -2.06 | 9.22 |  |  |  |
|  | Konica Minolta L* | -1.92 | -5.58 | 1.74 |  |  |  |
|  | Variable L* | 0.75 | -3.46 | 4.96 |  |  |  |
|  | Delfin Melanin Index | 3.06 | -1.82 | 7.94 |  |  |  |
|  | **Expected Total Effect<sup>1</sup>** | **-3.80** | -3.80 | -3.80 |  |  |  |

## Results

### Cohort Characteristics

From January 1, 2023 to June 30, 2023, a total of 1,167 admitted inpatients with qualifying SaO_2_-SpO_2_ pairs were screened at Duke University Hospital. Out of 301 patients who met our inclusion criteria and were approached, 134 patients consented to this study (see the flow diagram in Figure 1). After 6 exclusions due to withdrawal, missing location data, or incomplete data, 128 patients were considered for analysis (39.8% female, 43% Black, see Table 1).

**Figure 1.**
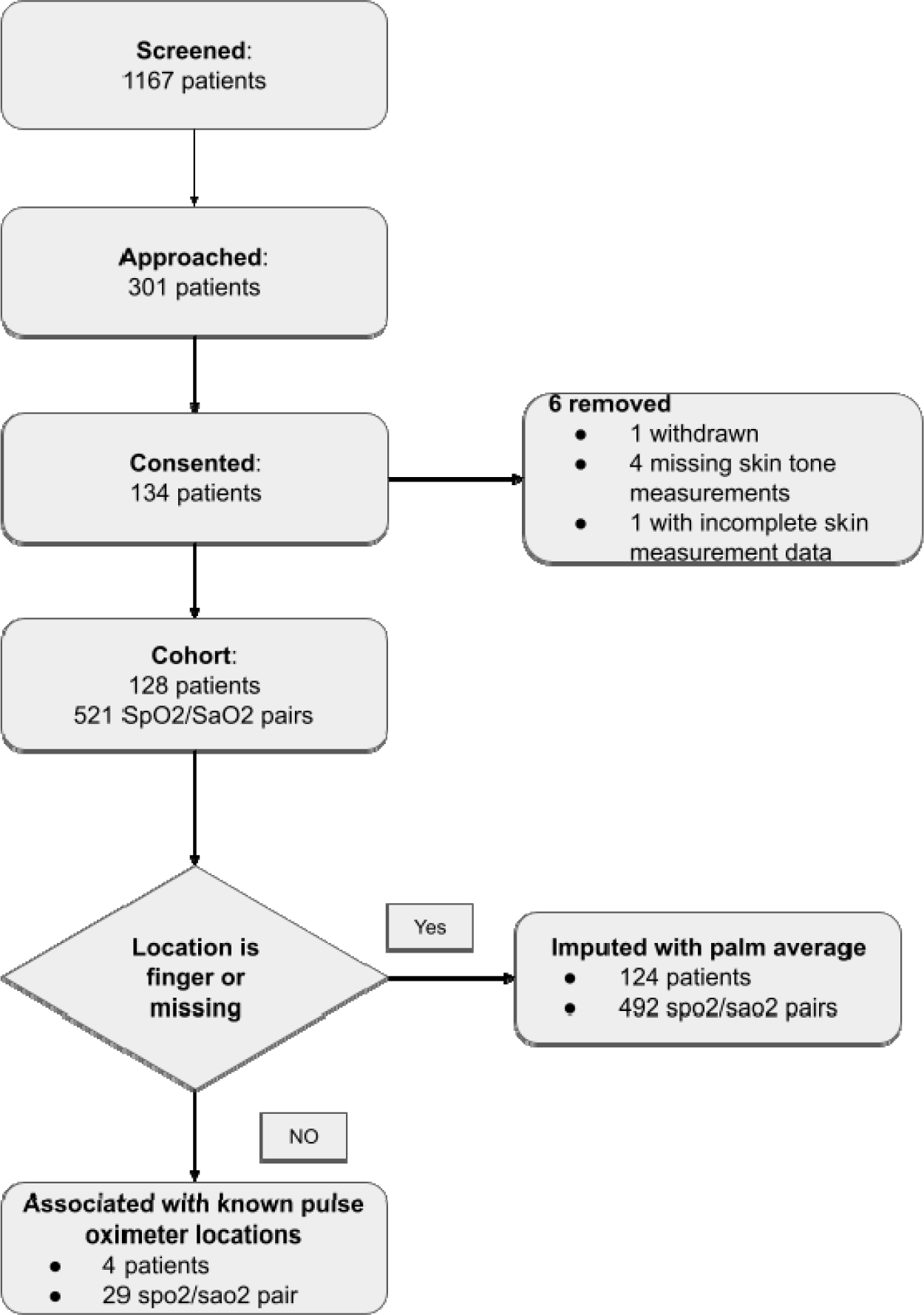
Flow diagram. A total of 1,167 patients were screened. Exclusion criteria included unremovable fingernail polish, admission for a vascular complication (e.g., grafting or stenting), amputation, and large areas of skin discoloration where the accuracy of skin tone measurements could be affected due to arterial insufficiency or cytopenias. Pairs containing either a SaO_2_ or a SpO_2_ measurement out of the 70–100% range were excluded.Of these, 301 patients qualified for this prospective study and were approached. Among the 134 patients who signed consent forms, one patient later withdrew, one patient didn’t have complete skin measurement data, and four patients didn’t have skin measurements. For patients who had pulse oximetry measurements done on the finger, we used the average of four palm locations (left ventral, right ventral, left dorsal, right dorsal). For patients who didn’t have pulse oximetry locations specified, we presumed the measurement was done on the finger and imputed it using the four palm locations as well.

After excluding readings that did not fall into the 70–100% range, a total of 521 SaO_2_–SpO_2_ pairs were obtained from this cohort. SpO_2_ values ranged from 82% to 100%, and SaO_2_ values ranged from 83.8% to 99.0%. The difference between SaO_2_ and SpO_2_ ranged from −9.0% to 8.8% – see Supplemental Table 1 for further pair-level characteristics.

### Measurement variability on skin tone scales

Supplemental Table 3 shows that objective scales resulted in lower standard deviations when compared to subjective scales. Palm averages are found to be more stable, when compared to other locations, supporting the design choice of taking palm averages as the preferred measurement for subsequent analyses.

### Skin tone and race data

Exploratory data analysis showed that the skin tone measurements yielded differences among each other and overlap with self-reported racial groups. Figure 2 depicts the unadjusted mean directional bias, variability of bias, and A_RMS_, per skin tone tertile. Further detailed text can be found in the Supplemental Text.

**Figure 2.**
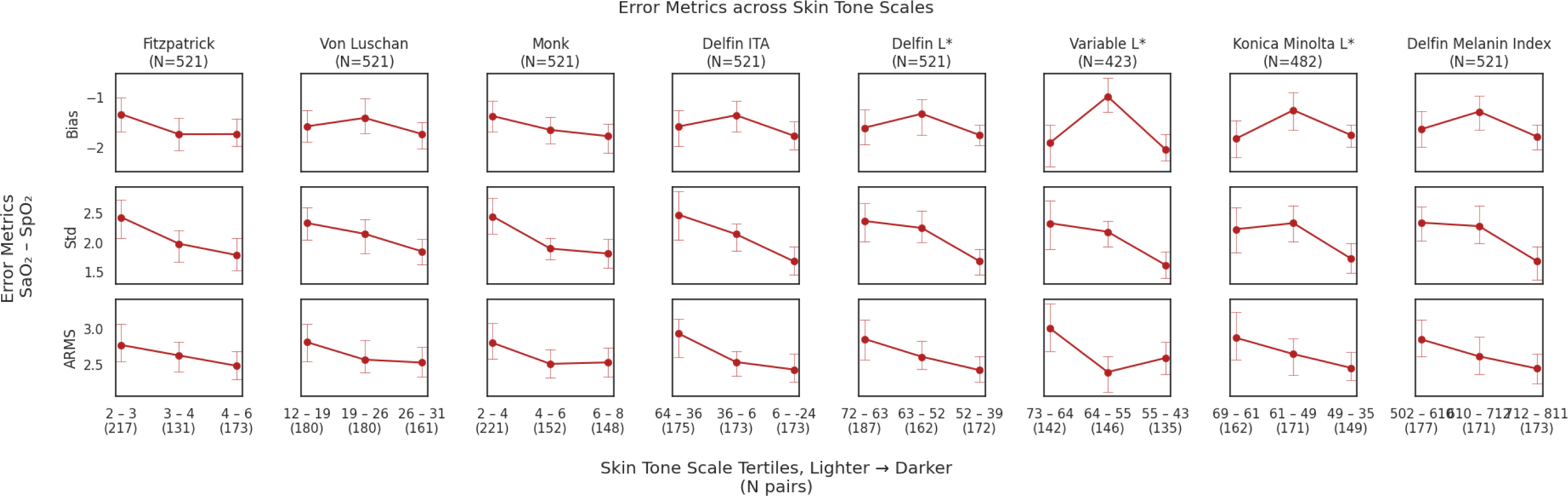
Unadjusted error metrics in pulse oximetry across skin tone scale tertiles. Unadjusted error metrics of mean directional bias, standard deviation, and accuracy root mean square (also known as A_RMS_ or root mean square error), across tertiles. Tertiles are ordered from lightest to darkest, from the left to the right on the x-axis. Note that a pulse oximetry bias defined as SaO_2_ - SpO_2_ results in a negative bias reflecting that pulse oximetry overestimates true oxygenation values. Fitzpatrick and Monk appear to have a trend towards more negative bias (e.g., bias increasingly negative) from lighter to darker tertiles. A_RMS_ appears to be lower (that is, a lower root mean square error) in many darker tertiles than in lighter tertiles. Variable L* and Konica Minolta L* have fewer patients because there was more missingness. Some patients did not have these measurements either due to patient refusal (often due to feelings of being overwhelmed, stress, or experiencing pain) or interruptions by clinical workflow.

### Linear mixed effect model on mean directional bias

To understand the effect of self-reported race and each of the skin tones on pulse oximetry discrepancy, the fitted linear mixed-effects model did not find race to have a significant effect (□2 = 0.90*, p* = 0.64), Nevertheless, the obtained coefficient is in the expected direction (- 0.23%, 95% CI: −0.76%, 0.30% for Black patients). Among eight skin tone measurements, only the Monk scale yielded a significant effect on the mean directional bias (point estimate: −2.40%; 95% CI: −4.32%, −0.48%; p-value: 0.01). – see Table 2 for a full report of the models.

To examine the combined effect of the six skin tone variables, excluding Konica Minolta L* and Variable L* due to missing data (the goal being to maximize sample size), the LRT (Supplemental Formula 7) rejected the null hypothesis (□2 = 14.98, *p* = 0.02). This suggests that at least one of these six skin tone measurements affects the mean directional bias. However, in a sensitivity analysis that included all eight skin tone variables, the LRT (Supplemental Formula 8) showed borderline insignificance (□2 = 14.86, *p* = 0.06)

## Discussion

The objective of this study was to investigate the relationship between skin tone measurements and pulse oximetry discrepancies. We prospectively collected skin tone data from 128 critically-ill patients, comprising a total of 521 pairs of pulse oximetry-ABG data, and leveraging six different tools across sixteen body sites. We addressed the fundamental limitation of previous studies on pulse oximetry racial discrepancies,^7–10, 12–14, 16^ which solely relied on racial or ethnic groups as a proxy for skin tone. As a prospective cohort study where we enrolled patients in a comprehensive screening process, we minimized assumptions associated with secondary data analysis^36^ and obtained a cohort with equal representation of Black and White patients.

The collected skin tone measurements were shown to yield differences among scales and when compared to self-reported race, suggesting that skin tone data carries information beyond self-reported race. When assessing the relation of skin tone with pulse oximetry bias, racial and ethnic disparities^7^ do seem to persist, whereby darker patients show a higher degree of mean directional bias (see Figure 2 and Table 2). These findings are aligned with a recent similar report.^37^ As opposed to a model that solely relies on self-reported race (and clinical confounders) to account for pulse oximetry bias – where the effect of self-reported race is found to be nonsignificant – models that accounted for skin tone found at least one of the measures to be significant (Supplemental Formula 7, results in Table 2). This finding suggests that skin tone is related, beyond self-reported race, to pulse oximetry bias.

In this study, we tested different devices for skin tone assessment that ranged from a negligible cost (color-printed scales) to thousands of dollars (Konica Minolta’s spectrophotometer). The measurement variability was non-negligible, but lower for objective scales, as expected. However, in exploring pulse oximetry bias, the Monk scale – designed to be easy-to-use for evaluation of technology, while representing a broad range of skin tones ^30, 38^ – was found to yield the strongest association with pulse oximetry bias (Table 2). As this study does not show clear evidence that more sophisticated and expensive devices (colorimetry or spectrophotometry) add value in this application of skin tone characterization, we believe that further investigation is necessary, including a broader range of skin tone measurement devices and a larger sample size.

In the exploratory analysis, both across self-reported race and skin tone measurements, darker-pigmented patients observed a lower SaO_2_–SpO_2_ variability (see Supplemental Table 4 and Figure 2). To assess precision while adjusting for confounding, we identified the within-*stratum* variation by modeling heterogeneous variance in the linear mixed-effects models (Supplemental Formula 9) using the tertiles defined in Figure 2. Supplemental Table 6 lists the built models, where the within-*stratum* variance is shown to be consistently lower in the darkest tertile of all eight skin tone measurements, and higher in the lightest tertile, except for Delfin Melanin Index. Consequently, the darker-pigmented patients of our cohort seem to have more consistently wrong pulse oximetry readings, despite having a lower A_RMS_. Although this finding is not aligned with a recent report,^37^ concerns about the reliability of A_RMS_ among different racial groups in pulse oximetry performance evaluation and potential inequitable treatment delay had already been raised before.^39^ Considering the far-reaching implications of these findings in pulse oximetry regulation, further investigation is necessary.

Considering the prohibitive cost of replacing existing pulse oximeters,^40^ our work stands as a fundamental milestone to any interim solution that may tackle pulse oximetry inaccuracies leveraging existing technologies. If the finding that pulse oximeters are “more consistently wrong” among darker-skinned patients stands in follow-up studies, one could argue that clinical algorithms that perform a holistic correction – and not simple race corrections^41^ – could be more attainable, due to the observed lower variance among darker-skinned patients. Given this finding, which suggests that skin tone may provide additional information beyond race, we propose that incorporating skin tone measurements could help mitigate residual confounding in algorithms solely reliant on self-reported race.

### Implications in regulation and pulse oximetry clearance

In response to FDA’s recent discussions on pulse oximetry performance discrepancies,^23–25^ we believe that this pilot study provides initial evidence to support the suggested need of thoughtfully collecting and assessing skin tone data in pulse oximetry clearances. Besides being an important factor in pulse oximetry miscalibration, and a more objective measure than self-reported race, skin tone data seems to yield utility in pulse oximetry discrepancies. Consequently, besides requiring racial and ethnic diversity for pulse oximetry clearance,^26^ we recommend the FDA to require the quantification and representation of a full spectrum of skin tones, while not disregarding the potential impact of other unmeasured confounders. Recognizing Beer-Lambert’s law’s impact on light transmission, we underline the importance of assessing other potential confounding variables such as perfusion, skin thickness, systemic vascular resistance, or local vascular resistance, for which further investigation is necessary as these are not commonly measured in medicine.

Moreover, our findings on increased variability of bias and better A_RMS_ among darker-skinned patients, despite worsened mean directional bias, raise questions about FDA’s conventional reliance on A_RMS_ for pulse oximetry clearance. These considerations require larger follow-up studies and are not necessarily aligned with other reports.^37^ Considering that bias in the direction of overestimation of SaO_2_ may carry more downstream clinical harm than bias in the opposite direction, we would like to build upon previous concerns^39^ and bring to debate the question: “What is an equitable performance assessment metric for pulse oximetry clearance?”.

### Limitations and Future Work

As our skin tone data presented non-negligible measurement variability across sites and examiners, we considered the average of the left and right, dorsal and ventral palm readings in 125 patients, out of 128, to obtain more stable skin tone measurements. Due to our limited sample size, most of the findings are not significant despite a reasonable effect size. In the future, we would like to run a larger study with more patients. Although this study’s cohort had over 40% Black patients, the darkest skin tones are rarely observed, which might be due to the population skin tone distribution of the community where our study was based. Moreover, we would like to enroll more patients with hypoxemia (i.e., SaO_2_ < 88%), to potentially investigate the impact of skin tone in hidden hypoxemia phenomena. Additionally, we would like to examine other potential covariates that may contribute to pulse oximetry disparities. Finally, as our prospective study suggests that, despite its effect, skin tone is unlikely to be the sole contributor to pulse oximetry discrepancies, we advocate for the need of further investigation on other unmeasured confounders.

## Conclusion

This pilot study analyzed skin tone measurements with pulse oximetry performance discrepancies among critically ill patients. We prospectively collected skin tone assessments via administered visual scales, reflectance colorimetric, and spectrophotometric devices. Pulse oximetry varied across skin pigmentation and, similarly to previous reports, darker-skin-toned patients yielded a greater bias, independently of clinical confounders. However, with a large variation in pulse oximetry data, skin tone is unlikely to be the sole contributor to performance discrepancies in pulse oximetry. While these findings necessitate a larger sample size to be further validated and select the best method(s) for skin tone measurement, we hope this paper provides a framework for future similar studies, as well as initial evidence to support FDA’s discussions on regulation changes towards more equitable pulse oximeters.

## Data Availability

An IRB revision is underway to evaluate this possibility but legal counsel has suggested that re-consent may be necessary for submission to a public access-controlled archive.

