## Supplementary material for "Utility of skin tone on pulse oximetry in critically ill patients: a prospective cohort study": ENCoDE - paper 1 - skin tone data characterization (Supplementary Material) - v1.0.docx

### Supplemental Formulas

#### Supplemental Formula 1. Normalization of skin tone distributions

Normalized distributions of skin tone data by each measure are demonstrated in Supplemental Figure 1. This is further extended for Black (N = 57) and White (N = 57), in Supplemental Figure 2. In these distributions, the following formula, Supplemental Formula 1 is used:

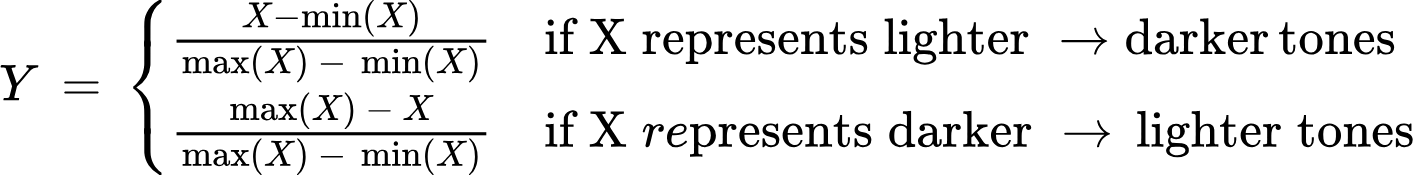

where Y represents the normalized distribution and X is the original distribution, according to the skin tone scales.

#### Supplemental Formula 2. Arterial blood gas (SaO_2_)–pulse oximetry (SpO_2_) mean directional bias

We defined the measurement bias of pulse oximetry (SpO_2_) as the value of SaO_2_ - SpO_2_ throughout the whole manuscript and supplementary. The mean directional bias is the average of the measurement bias represented in the formula below.

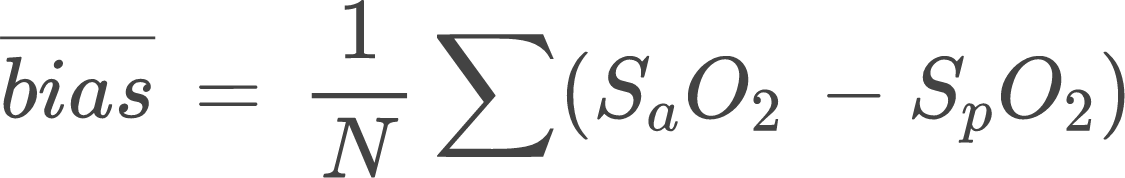

#### Supplemental Formula 3. Variation of bias (SaO_2_–SpO_2_ Precision)

We defined variation of bias as the standard deviation of the difference between SpO_2_ and SaO_2_.

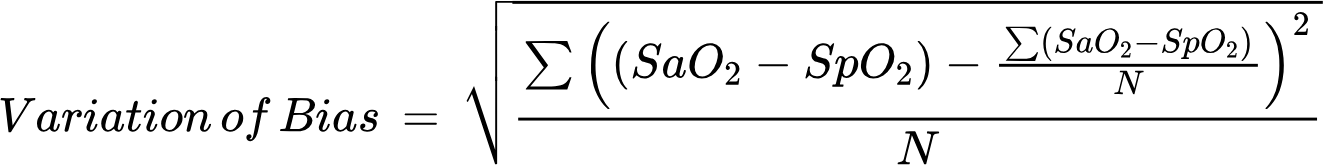

#### Supplemental Formula 4. SaO_2_–SpO_2_ accuracy root mean square (A_RMS_)

A_RMS_ can be derived from both the mean directional bias and precision, and in conformance with Clause 201.12.1.101.1 of ISO 80601-2-61:2011.^27^ In 2013, the FDA issued a guidance document to assist industry in preparing premarket notifications (510(k)s) for pulse oximeters. According to the FDA, transmittance, wrap and clip pulse oximeters must meet a clearance threshold of A_RMS_ ≤ 3.0% between SaO_2_ and SpO_2_ measurements ranging from 70–100%.^27^

A_RMS_ is equivalent to root mean square error (RMSE).

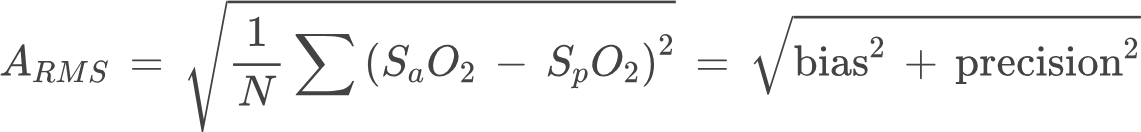

#### Supplemental Formula 5: Linear mixed-effects model with self-reported race

Linear mixed-effects model with random effect of individual patients (Record_ID_) demonstrating the fixed effect of race on the measurement bias of SpO_2_, with clinical variables adjusted, clinical variables contain heart rate, mean arterial pressure (MAP), SaO_2_, and blood pH levels.

Full model:

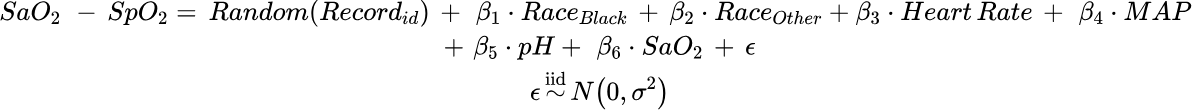

Reduced model:

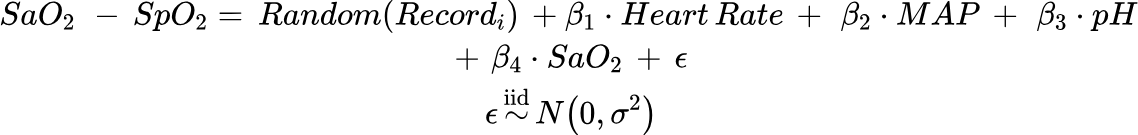

Hypothesis testing:

The null hypothesis (H_0_) implies that with self-reported race added the goodness of fit does not differ from the reduced model. The alternative hypothesis implies with race added the goodness of fit is significantly different from the reduced model.

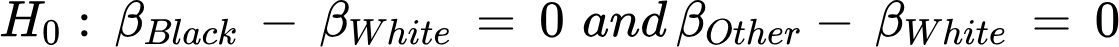

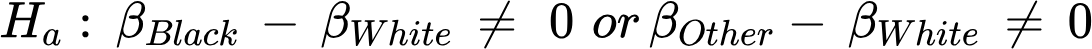

#### Supplemental Formula 6: Linear mixed-effects models with separate continuous skin tone measurements

Linear mixed-effects models with random effect of individual patients (Record_ID_) demonstrating the effect of single skin tone measurements on the measurement bias of SpO_2_, with Race and clinical variables adjusted, clinical variables contain heart rate, mean arterial pressure (MAP), SaO_2_, and blood pH levels. Modeling and hypothesis testing were performed for each of the Skin tones. Skin tone scale includes Monk, Fitzpatrick, VonLuschan, Delfin Melanin Index, Delfin
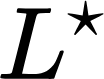
, Konica Minolta
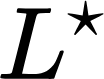
, Variable
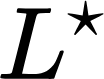
, and Delfin ITA.

Full model:

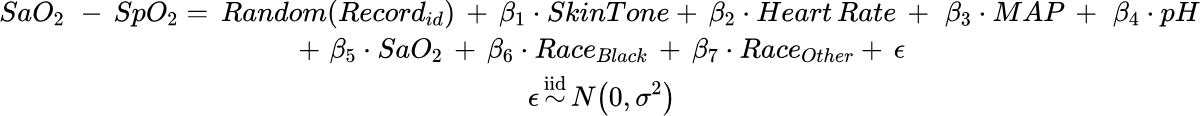

Reduced model:

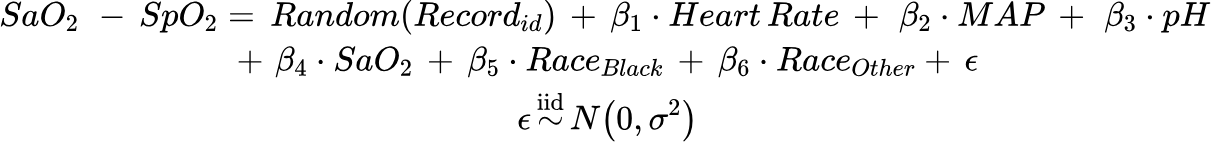

Hypothesis testing:

The null hypothesis (H_0_) implies that with each single skin tone scale added the goodness of fit does not differ from the reduced model. The alternative hypothesis implies with each single skin tone scale added the goodness of fit is significantly different from the reduced model.

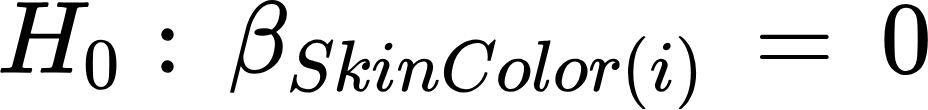

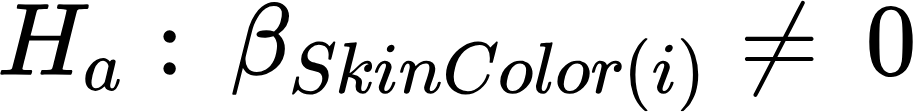

#### Supplemental Formula 7: Single linear mixed-effects model with skin tone measurements together that do not have missingness

Linear mixed-effects models with random effect of individual patients (Record_ID_) demonstrating the effect of six skin tone measurements on the measurement bias of SpO_2_, with race and clinical variables adjusted.

Full model:

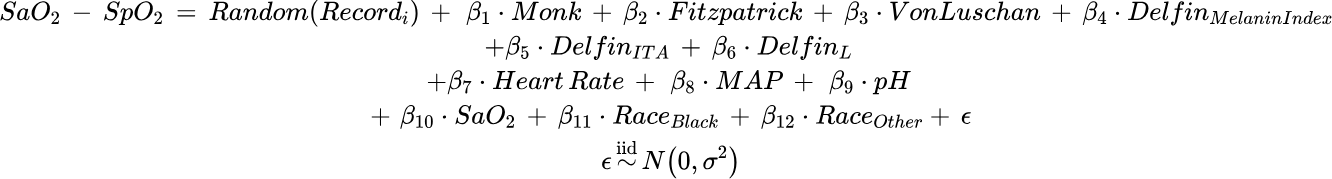

Reduced model:

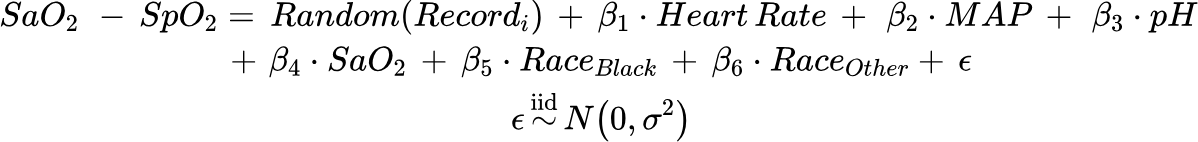

Hypothesis testing:

The null hypothesis (H_0_) implies that with skin tone measurements added the goodness of fit does not differ from the reduced model. The alternative hypothesis implies with skin tone measurements added the goodness of fit is significantly different from the reduced model.

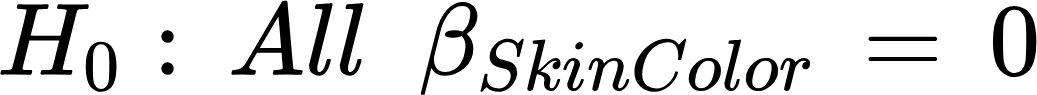

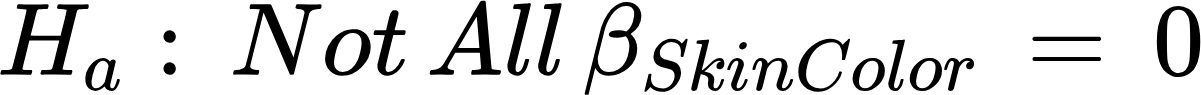

#### Supplemental Formula 8: Single linear mixed-effects model with all eight skin tone measurements together (sensitivity analysis)

Linear mixed-effects model with random effect of individual patients (Record_ID_) demonstrating the effect of all eight skin tone measurements on the measurement bias of SpO_2_, with Race and clinical variables adjusted.

Full Model:

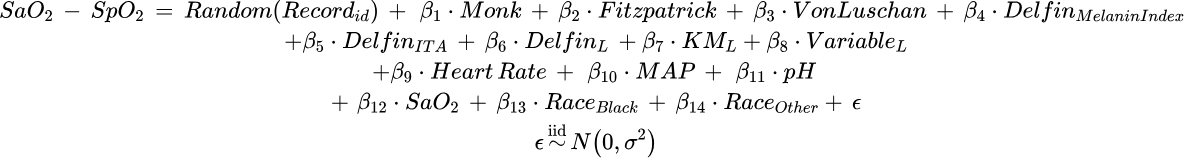

Reduced Model:

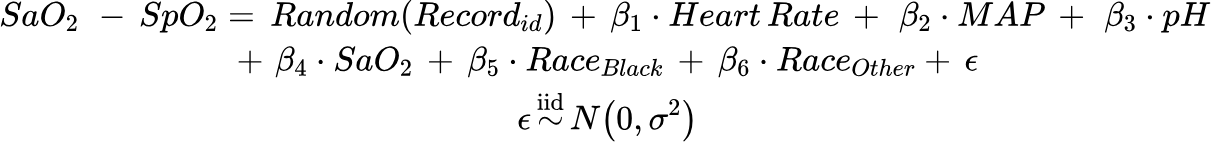

Hypothesis testing:

The null hypothesis (H_0_) implies that with skin tone measurements added the goodness of fit does not differ from the reduced model. The alternative hypothesis implies with skin tone measurements added the goodness of fit is significantly different from the reduced model.

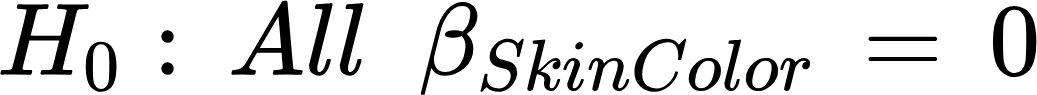

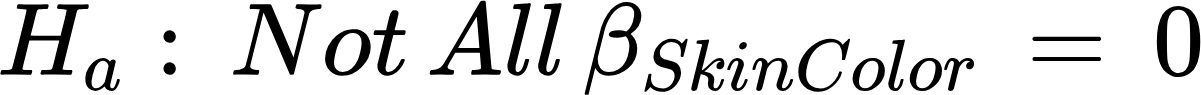

#### Supplemental Formula 9: Linear mixed-effects models with separate skin tone measurements, with heterogeneous variance on tertile of skin tone

Full model:

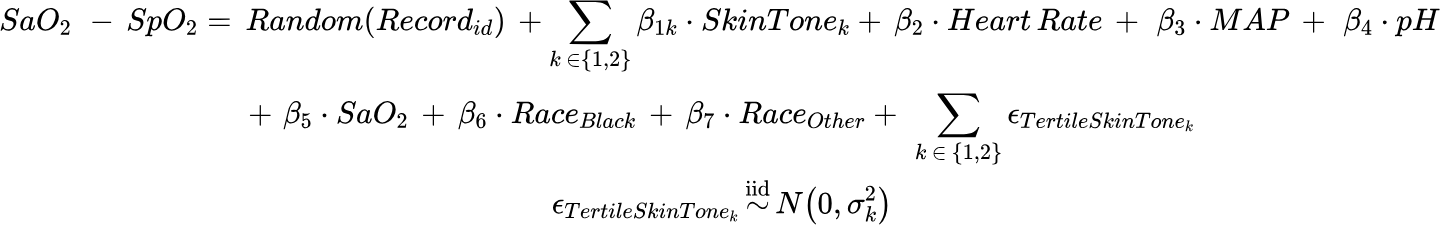

Reduced model:

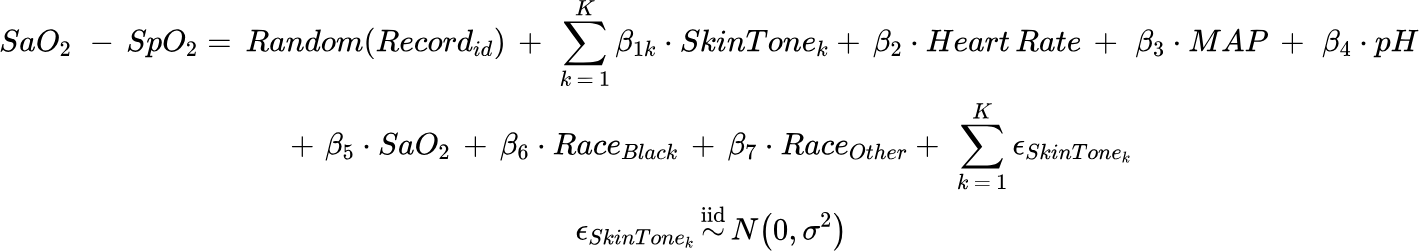

Linear mixed-effects models with random effect of individual patients (Record_ID_) and heterogenous variation fitted across skin tone tertile investigating variabilities of bias across each of the eight skin tone scales, with Race and clinical variables adjusted. Implemented with the “VarIdent” function in the “nlme” package.

Skin tone scales include Monk, Fitzpatrick, VonLuschan, Delfin Melanin Index, Delfin
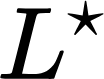
, Konica Minolta
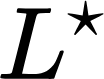
, Variable
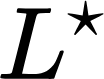
, and Delfin ITA. Tertiles (k) include light, medium, and dark.

Hypothesis testing:

The null hypothesis (H_0_) implies that fitting each skin tone tertile with heterogeneous variance added to the goodness of fit does not differ from the reduced model. The alternative hypothesis implies with heterogeneous added the goodness of fit is significantly different from the reduced model.

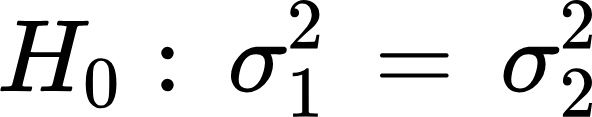

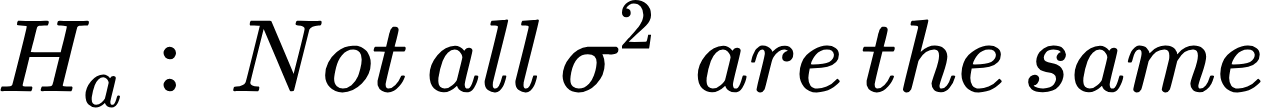

### Supplemental Text

#### Supplemental Methods

##### Skin tone data

Skin tone measurements were taken at sixteen different locations: 8 in the left and right upper extremities (dorsal and ventral finger pad, dorsal and ventral palm); 3 on the head (forehead, inner and outer surface of an earlobe); 1 on the sternum, and 4 on the left and right lower extremities (dorsal and ventral toe).

Although all these locations were recorded, the skin tone measurement used in subsequent analyses was the average of 4 locations from patients' palms (left dorsal, right dorsal, left ventral, right ventral). Palm measurements were preferred over finger location because fingers are subject to high variability, while the average of 4 palm locations is a more stable measurement (see Supplemental Table 3).

For comparability, skin tone distributions are then normalized across the minimum and maximum observed values for each measure, to be in a unitary range, as defined in Supplemental Formula 1.

We have quantified the measurement variability of the eight skin tone measures for a single, external subject, across 3 different days, 3 different examiners, and 18 locations (the sixteen aforementioned locations plus the left and right under arm). Similarly to the cohort’s skin tone measurements, the skin tone of this external subject was normalized using Supplemental Formula 1, but the minimum and maximum values correspond to the absolute lower and upper bounds of the scales, as defined in Supplemental Table 3.

##### Standardization of skin tone data collection

Timing of measurements and standardizations of the clinical environment is a critical aspect of data collection that is commonly overlooked. In particular, an inpatient population presents complications when attempting to achieve a more fixed environment for assessments. Complications can potentially be improved by timely data acquisition, understanding the unique workflow of the care team and current patient trajectory, and adjusting each patient’s room to represent a standardized environment.

This study required a necessity of data-time synchronization to minimize potential discrepancies between synchronized ABG-pulse oximetry measurements and skin tone assessments. To account for feasibility, skin tone assessments were performed within 7 days of SaO_2_ SpO_2_ pairs to enable a clearer comprehension of the patient’s stable state. Due to the criticality of the patients, assessments were collected from patients lying down or in a seated position.

Additionally, assessments were performed in patient rooms of the ICU as well as floor units in the event that the patient transferred out of the ICU at any point in time during their enrollment. Variations of lighting alone have the ability to skew color perception which can lead to misclassification of skin tone due to appearing lighter or darker during assessment. To account for the unpredictability and dynamic environment of the ICU, attempts to control lighting were made by shutting all blinds and turning on all light sources surrounding the patient's bed. For earlobes (and fingers/toes if needed), a black card was placed on the opposite side to reduce the impact of reflected light. Given the difficulty in finger data measurement due to shape and size, skin tone data was averaged between dorsal and ventral palms for both hands.

##### Exploratory data analysis

Analyses were performed in Python 3.10.

###### Patient and SaO_2_–SpO_2_ pairs characteristics

Python’s *tableone* package^32^ was used to compute summary statistics of the obtained cohort, both at patient and SaO_2_–SpO_2_ pair-level. Means and standard deviations were computed across racial groups, as shown in [Table 1](https://docs.google.com/document/d/13LajNHsU56uFxhYAwGn_eaPTR0eB8q3G_3orbXuHSfk/edit#heading=h.1tuee74) and Supplemental Table 1.

###### Measurement variability

The standard deviation was computed across the different values of the same measure and location, and compared with the average standard deviations across all locations. As a sanity check of our design choice, average standard deviations across palm and finger locations were also computed, as depicted in Supplemental Table 3.

###### Skin tone distributions and concordance across scales

After normalization of skin tone measurements, distributions were compared using kernel density estimation plots in the format of ridge plots with the different skin tone scales in parallel. As a sanity check, these distributions were performed for the whole dataset (Supplemental Figure 2) and separating Black from White patients (Supplemental Figures 3a and 3a, respectively). Additionally, concordance across scales was assessed by comparing each skin tone variable against each other. A triangular matrix of 2D hexagonal binning plots (each set to have 10 full hexagons on the x-axis) is reported in Supplemental Figure 1.

###### Skin tone tertiles and race groups

Each normalized skin tone measurement was discretized into tertiles such that each bin was (approximately) equally sized, with the goal of separating lighter from darker patients (which would be in the first and third tertile, respectively, as compared to the median darkness in the middle tertile). To assess the extent to which race and skin tone overlap, a cross-tabulation of both covariates was performed and displayed in the form of heatmaps (Supplemental Figure 4).

###### Pulse oximetry performance across skin tone tertiles

For each tertile, mean directional bias, variability of bias, and A_RMS_ were computed. To provide an estimate of uncertainty, 95% confidence intervals (CI) are obtained via a 100-iteration bootstrap resampling of each race subgroup. For each resampling iteration, the three error metrics were computed and the 2.5% and 97.5% percentiles were then considered as the 95% CI. Finally, this was reported in a matrix of plots ([Figure 2](https://docs.google.com/document/d/13LajNHsU56uFxhYAwGn_eaPTR0eB8q3G_3orbXuHSfk/edit#heading=h.pas1xy95yn5)), as well as in Supplemental Table 5, containing further details of the obtained estimations.

A similar methodology was followed across race groups (Black and White patients only) to assess whether previous literature findings on racial bias were confirmed using our data (Supplemental Table 4).

#### Supplemental Results

##### Skin tone data

The first step in analyzing the collected skin tone is to assess the magnitude of measurement variability which, in the scope of this study, consisted of evaluating the consistency of the same measurement of a single, external subject across three different examiners and three different days. Supplemental Table 3 shows that objective scales result in lower standard deviations, when compared to subjective scales, as expected. Among the subjective scales, Von Luschan’s scale seems to yield the lowest measurement variability, whereas Variable L* appears to be the one yielding the lowest error among the objective scales. When comparing the different measurement sites, palms, sternum, and under arm exhibit lower errors. Finally, palm averages are found to be more stable and less prone to errors, when compared to other locations, supporting the design choice of taking palm averages as the preferred measurement for subsequent analyses.

To assess the consistency of skin tone representation across scales, pairwise skin tone variable comparisons are reported in Supplemental Figure 1. Specifically, while most variables present linear relations, with an identity relation being observed, Delfin Melanin Index seems to have a sigmoidal association with the remaining variables, while Konica Minolta L* seems to present quadratic relations.

##### Skin tone data and race

Such variations in describing skin tone are further confirmed when comparing the normalized distributions of the different skin tone scales. Supplement Figure 2 shows that the eight skin tone variables assume different normalized distributions that are not necessarily concordant among each other. These variations persist when separating Black (Supplemental Figure 3a) and White (Supplemental Figure [Supplemental Figure 3](https://docs.google.com/document/d/13LajNHsU56uFxhYAwGn_eaPTR0eB8q3G_3orbXuHSfk/edit#heading=h.kgcv8k)b) patients. Reassuringly, splitting by race group skews the distributions to either side, although overlap persists to a considerable extent, indicating that patients with different self-reported races may have similar skin tones.

When we divide our cohort into tertiles based on skin tone, the darkest tertile consists mostly of Black patients, and the lightest tertile consists mostly of White patients (see Supplemental Figure 4). However, the middle tertile of the eight skin tone measurements often includes a combination of patients across different racial groups, which indicates that patients from different race groups and skin tones overlap and that skin tone data contains information that goes beyond self-reported race.

##### Unadjusted mean directional bias, variability of bias, and A_RMS_ across race

As a baseline that is comparable with literature’s previous reports on racial bias, Supplemental Table 4 depicts three considered error metrics computed across racial groups. Similarly to literature, mean directional bias seems to be higher, in absolute terms, among Black patients (i.e., increased chance of SaO_2_ overestimation). Due to our limited sample size, these differences are not significant, with a mean directional bias of -1.618, 95% CI: (-1.857, -1.467) in Black patients, versus -1.494, 95% CI: (-1.735, -1.263) in White patients. Additionally, and contrary to previous findings in literature, Black patients yield a lower pulse oximetry–ABG standard deviation of 1.876, 95% CI: (1.681, 2.036), as compared to White patients 2.370, 95% CI: (2.089, 2.641). This opposite trend in precision results in equally unexpected trends with A_RMS_, which is also lower (although not significant) among Black patients. Finally, it is worth noting that the expected values of A_RMS_, including the upper bounds of the computed 95% CIs, are below the clearance threshold imposed by FDA, of 3.0%.

##### Unadjusted mean directional bias, variability of bias, and A_RMS_ across skin tone tertiles

When performing a similar analysis across skin tone tertiles, the directionality of the aforementioned findings stands in all eight skin tone variables, when comparing the lighter with the darkest tertile, although not significantly different. Specifically, the darker the patient’s skin tone, the worse the mean directional bias, and the better the precision is, resulting in a better A_RMS_ – see [Figure 2](https://docs.google.com/document/d/13LajNHsU56uFxhYAwGn_eaPTR0eB8q3G_3orbXuHSfk/edit#heading=h.pas1xy95yn5) for an overview of these trends, and Supplemental Table 4 for the expected values with three decimal points.

Furthermore, our results show that mean directional bias is not uniformly distributed across the three tertile skin tones. Darker skin tone measurements are associated with a linear worsening of bias (i.e., more negative values, thus a higher chance of SaO_2_ overestimation) when using the Fitzpatrick and Monk scales. However, bias is not linearly associated with skin tone among the rest of the skin tone measurements. This could be due to the fact that the middle tertile contains patients with different self-reported races, as reported previously in Supplemental Figure 4.

On the other hand, the standard deviation consistently decreases as skin tone gets darker, which translates to a higher precision among dark-skinned patients. Since the precision increase is more pronounced than the decrease of mean directional bias, as we go from lighter to darker-skinned patients, the resulting average A_RMS_ is reported to be lower among darker skin tones. These differences in A_RMS_ are consistent in all skin tone measurement scales, except for Variable L*.

Supplemental Figure 5 shows the mean directional bias across SpO_2_ ranges and Monk scale’s tertiles.

### Supplemental Tables

#### Supplemental Table 1. Characteristics of arterial blood gas samples and paired pulse oximetry by race group.

This table depicts the pair-level characteristics of the obtained cohort, in terms of sex distribution, pulse oximetry and ABG measurements, as well as paired laboratory test values. The latter present missingness due to the applied pairing criteria; nevertheless, covariates’ missingness is always below 10% (in the 521 pairs).

Abbreviations: MAP, main arterial pressure. BUN, blood urea nitrogen.WBC, white blood cell.

|  |  | **SaO2-SpO2 pairs grouped by race group** | | | |
| --- | --- | --- | --- | --- | --- |
|  | **Missing** | **Black** | **Other** | **White** | **Overall** |
| **n** |  | 232 | 24 | 265 | 521 |
| **Sex (Female), n (%)** | 0 | 91 (39.2) | 3 (12.5) | 107 (40.4) | 201 (38.6) |
| **SaO_2_ (%), mean (SD)** | 0 | 95.6 (2.4) | 95.9 (2.0) | 95.8 (2.2) | 95.7 (2.3) |
| **SpO_2_ (%), mean (SD)** | 0 | 97.2 (3.1) | 97.7 (2.2) | 97.3 (2.9) | 97.3 (2.9) |
| **SaO_2_ – SpO_2_ (%), mean (SD)** | 0 | -1.6 (1.9) | -1.8 (1.8) | -1.5 (2.4) | -1.6 (2.1) |
| **pH, mean (SD)** | 39 | 7.4 (0.1) | 7.4 (0.1) | 7.4 (0.1) | 7.4 (0.1) |
| **Heart rate (BPM), mean (SD)** | 8 | 93.5 (18.9) | 94.1 (15.5) | 96.1 (19.6) | 94.8 (19.1) |
| **MAP (mmHg), mean (SD)** | 19 | 81.7 (17.9) | 95.8 (54.1) | 81.4 (33.5) | 82.2 (29.3) |
| **Sodium (mmol/L), mean (SD)** | 15 | 139.0 (5.3) | 138.2 (2.7) | 137.5 (5.7) | 138.2 (5.4) |
| **Platelet (10^9^/L), mean (SD)** | 28 | 180.1 (102.5) | 279.8 (186.4) | 202.2 (125.8) | 195.9 (121.3) |
| **Potassium (mmol/L), mean (SD)** | 14 | 4.1 (0.6) | 3.9 (0.6) | 4.0 (0.5) | 4.1 (0.6) |
| **BUN (mg/dL), mean (SD)** | 26 | 24.3 (17.4) | 20.2 (11.0) | 31.8 (22.7) | 27.9 (20.4) |
| **WBC (10^9^/L), mean (SD)** | 44 | 13.9 (7.1) | 17.2 (7.6) | 14.5 (8.7) | 14.4 (8.0) |
| **Creatinine (mg/dL), mean (SD)** | 26 | 1.5 (1.0) | 1.5 (1.4) | 1.9 (1.4) | 1.7 (1.3) |
| **Glucose (mg/dL), mean (SD)** | 9 | 152.7 (54.1) | 138.3 (27.6) | 151.6 (58.4) | 151.5 (55.4) |
| **Bicarbonate (mg/dL), mean (SD)** | 26 | 24.7 (3.8) | 23.7 (3.0) | 23.7 (5.0) | 24.1 (4.4) |
| **Chloride (mmol/L), mean (SD)** | 26 | 105.3 (6.1) | 105.5 (4.4) | 103.8 (6.5) | 104.6 (6.3) |
| **Hemoglobin (g/L), mean (SD)** | 15 | 10.0 (1.9) | 11.2 (1.3) | 9.5 (1.6) | 9.8 (1.8) |
| **Norepinephrine equivalent dose (mcg/kg/min), mean (SD)** | 0 | 0.1 (0.1) | 0.0 (0.0) | 0.1 (0.1) | 0.1 (0.1) |

#### Supplemental Table 2. Skin tone variables and respective used devices and characteristics

The eight skin tone measurements, which can be split into three types – administered visual scales, reflectance colorimetry, and reflectance spectrophotometry – are further explored in this table. The direction of skin tone characterization is reported (lower values correspond to lighter patients or the other way around), as well as the measurement bounds, minimum and maximum values, and device names. Finally, we provide a relative cost, from negligible to lowest, medium and highest.

| **Type** | **Skin Tone Variable** | **Direction** | **Scale Lower Bound** | **Scale Upper Bound** | **Minimum palm average** | **Maximum palm average** | **Device** | **Cost** |
| --- | --- | --- | --- | --- | --- | --- | --- | --- |
| Administered visual scales **(subjective)** | Fitzpatrick | Lighter → Darker | 1 | 6 | 2 | 5.5 | Colored printed scale | Negligible |
|  | Von Luschan | Lighter → Darker | 1 | 36 | 12 | 31.25 |  |  |
|  | Monk | Lighter → Darker | 1 | 10 | 2 | 7.75 |  |  |
| Reflectance colorimetry **(objective)** | Delfin ITA | Darker → Lighter | -90 | 90 | -24.25 | 63.75 | SkinColorCatch | Medium |
|  | Delfin L* | Darker → Lighter | 0 | 100 | 38.75 | 71.5 |  |  |
|  | Delfin Melanin Index | Lighter → Darker | 0 | 999 | 502 | 810.75 |  |  |
| Reflectance spectrophotometry **(objective)** | Variable L* | Darker → Lighter | 0 | 100 | 42.87 | 72.51 | Variable Spectro 1  Pro Bridge Set | Lowest |
|  | Konica Minolta L* | Darker → Lighter | 0 | 100 | 35 | 69.07 | Konica Minolta CM-700D Spectrophotometer | Highest |

#### Supplemental Table 3. Skin tone measurement standard deviations in an external subject, across different locations, scales and averaging scenarios.

| **Location \ Scale** | **Fitzpatrick** | **Von Luschan** | **Monk** | **Delfin ITA** | **Delfin L*** | **Variable L*** | **Konica Minolta L*** | **Delfin Melanin Index** |
| --- | --- | --- | --- | --- | --- | --- | --- | --- |
| L finger dorsal | 0.219 | 0.119 | 0.130 | 0.123 | 0.050 | 0.036 | 0.077 | 0.037 |
| L finger ventral | 0.207 | 0.085 | 0.115 | 0.035 | 0.016 | 0.070 | 0.032 | 0.021 |
| L palm dorsal | 0.103 | 0.043 | 0.057 | 0.011 | 0.010 | 0.006 | 0.020 | 0.071 |
| L palm ventral | 0.151 | 0.076 | 0.115 | 0.023 | 0.019 | 0.015 | 0.022 | 0.024 |
| R finger dorsal | 0.151 | 0.116 | 0.122 | 0.184 | 0.061 | 0.053 | 0.213 | 0.044 |
| R finger ventral | 0.151 | 0.111 | 0.135 | 0.029 | 0.020 | 0.013 | 0.236 | 0.016 |
| R palm dorsal | 0.103 | 0.049 | 0.084 | 0.005 | 0.005 | 0.006 | 0.030 | 0.008 |
| R palm ventral | 0.151 | 0.069 | 0.091 | 0.017 | 0.017 | 0.016 | 0.029 | 0.014 |
| forehead | 0.103 | 0.035 | 0.130 | 0.007 | 0.008 | 0.013 | 0.042 | 0.008 |
| R ear lobe outer | 0.151 | 0.091 | 0.122 | 0.043 | 0.017 | 0.031 | 0.057 | 0.035 |
| R ear lobe inner | 0.151 | 0.082 | 0.141 | 0.128 | 0.086 | 0.047 | 0.176 | 0.089 |
| sternum | 0.103 | 0.087 | 0.070 | 0.007 | 0.012 | 0.016 | 0.045 | 0.006 |
| L toe dorsal | 0.301 | 0.113 | 0.093 | 0.088 | 0.023 | 0.048 | 0.069 | 0.018 |
| L toe ventral | 0.253 | 0.091 | 0.167 | 0.044 | 0.034 | 0.027 | 0.034 | 0.087 |
| R toe dorsal | 0.266 | 0.113 | 0.093 | 0.049 | 0.022 | 0.061 | 0.065 | 0.077 |
| R toe ventral | 0.197 | 0.091 | 0.148 | 0.070 | 0.038 | 0.031 | 0.017 | 0.044 |
| L under arm | 0.110 | 0.000 | 0.057 | 0.016 | 0.071 | 0.013 | 0.010 | 0.081 |
| R under arm | 0.110 | 0.000 | 0.057 | 0.020 | 0.080 | 0.008 | 0.011 | 0.041 |
| **Averages** |  |  |  |  |  |  |  |  |
| L + R, dorsal + ventral palms | 0.117 | 0.049 | 0.070 | 0.009 | 0.009 | 0.008 | 0.017 | 0.017 |
| L dorsal + ventral palm | 0.117 | 0.043 | 0.076 | 0.016 | 0.013 | 0.009 | 0.014 | 0.034 |
| R dorsal + ventral palm | 0.117 | 0.058 | 0.082 | 0.009 | 0.009 | 0.010 | 0.028 | 0.009 |
| L + R  dorsal + ventral fingers | 0.160 | 0.094 | 0.108 | 0.075 | 0.027 | 0.021 | 0.098 | 0.022 |
| L dorsal + ventral finger | 0.197 | 0.092 | 0.108 | 0.071 | 0.029 | 0.026 | 0.044 | 0.023 |
| R dorsal + ventral finger | 0.126 | 0.100 | 0.115 | 0.105 | 0.037 | 0.029 | 0.224 | 0.029 |
| **Overall Average** | **0.165** | **0.076** | **0.107** | **0.050** | **0.033** | **0.028** | **0.066** | **0.040** |

Each scale was normalized to be in range [0, 1], using the lower and upper possible bounds of each scale, available in Supplemental Table 3. The color scale represents the degree of variability: red cells correspond to higher values of standard deviation and, therefore, lower bounds; green cells correspond to higher variability; white cells correspond to the midpoint. The shading is then proportional to the variability degree, with the darkest green representing the maximum variability (i.e., minimum standard deviation), and vice versa for the darkest red cells. We note that the combination of L + R, dorsal + ventral palms presents the lowest variation, when compared to other sites.

#### Supplemental Table 4. Unadjusted mean directional bias, standard deviation, and A_RMS_ across Black and White patients.

Expected values are provided, as well as 95% confidence intervals (CI) that are obtained via a 100-iteration bootstrap resampling of each race subgroup (for each resampling iteration, the bias, standard deviation, and A_RMS_ are computed and the 2.5% and 97.5% percentiles are considered as the 95% confidence interval). Units are percentage (%), in terms of arterial oxygen saturation. The mean directional bias is lower among Black patients, while the precision and A_RMS_ are higher among Black patients, as compared to White patients.

| **Mean Directional Bias (95% CI)** | | **Precision (95% CI)** | | **A_RMS_ (95% CI)** | |
| --- | --- | --- | --- | --- | --- |
| **Black** | **White** | **Black** | **White** | **Black** | **White** |
| -1.618 (-1.857, -1.467) | -1.494 (-1.735, -1.263) | 1.876 (1.681, 2.036) | 2.370 (2.089, 2.641) | 2.477 (2.317, 2.655) | 2.802 (2.601, 2.987) |

#### Supplemental Table 5. Detailed Error Metrics across Skin Tone Scales’ tertiles

Detailed results, as shown in [Figure 2](https://docs.google.com/document/d/13LajNHsU56uFxhYAwGn_eaPTR0eB8q3G_3orbXuHSfk/edit#heading=h.pas1xy95yn5). Unadjusted error metrics of mean directional bias, standard deviation, and A_RMS_, across skin tone tertiles. Tertiles are ordered from lightest to darkest, from the left to the right, under each metric. For example, for the Monk skin tone scale, the mean directional bias is: lightest tertile: -1.371, 95% CI: (-1.646, -1.113); mid tertile: -1.643, 95% CI (-1.890, -1.340); darkest tertile -1.767, 95% CI: (-2.038, -1.515). Other skin tone scales show similar trends, whereby lightest tertiles present a lower mean directional bias. For precision and A_RMS_ the trends are reversed, with darker tertiles presenting lower precision and A_RMS_.

| **Error Metric** | **Mean Directional Bias (95% CI)** | | | **Precision (95% CI)** | | | **A_RMS_ (95% CI)** | | |
| --- | --- | --- | --- | --- | --- | --- | --- | --- | --- |
| **Skin Tone Scale** | **0% - 33.3%** | **33.3% - 66.6%** | **66.6% - 100%** | **0% - 33.3%** | **33.3% - 66.6%** | **66.6% - 100%** | **0% - 33.3%** | **33.3% - 66.6%** | **66.6% - 100%** |
| **Fitzpatrick** | -1.334  (-1.635, -1.054) | -1.727  (-2.057, -1.351) | -1.726  (-1.981, -1.462) | 2.438 (2.051, 2.824) | 1.989 (1.732, 2.263) | 1.794 (1.533, 2.057) | 2.779 (2.539, 3.045) | 2.634 (2.421, 2.894) | 2.489 (2.291, 2.668) |
| **Von Luschan** | -1.573 (-1.962, -1.188) | -1.408 (-1.752, -1.014) | -1.725 (-2.046, -1.426) | 2.339 (2.051, 2.631) | 2.156 (1.819, 2.397) | 1.855 (1.577, 2.100) | 2.819 (2.542, 3.057) | 2.575 (2.332, 2.770) | 2.534 (2.329, 2.758) |
| **Monk** | -1.371 (-1.646, -1.113) | -1.643 (-1.890, -1.340) | -1.767 (-2.038, -1.515) | 2.451 (2.116, 2.739) | 1.905 (1.677, 2.143) | 1.820 (1.600, 2.066) | 2.808 (2.604, 3.094) | 2.516 (2.331, 2.721) | 2.536 (2.317, 2.798) |
| **Delfin ITA** | -1.577 (-1.951, -1.190) | -1.355 (-1.607, -1.088) | -1.758 (-2.004, -1.532) | 2.481 (2.131, 2.842) | 2.150 (1.872, 2.417) | 1.689 (1.433, 1.914) | 2.939 (2.632, 3.206) | 2.541 (2.324, 2.749) | 2.438 (2.271, 2.645) |
| **Delfin L*** | -1.602 (-1.864, -1.242) | -1.325 (-1.642, -0.965) | -1.745 (-2.015, -1.507) | 2.374 (2.021, 2.721) | 2.255 (1.992, 2.486) | 1.690 (1.463, 1.896) | 2.864 (2.506, 3.118) | 2.616 (2.420, 2.790) | 2.429 (2.244, 2.629) |
| **Variable L*** | -1.897 (-2.235, -1.576) | -0.986 (-1.276, -0.702) | -2.024 (-2.325, -1.787) | 2.336 (1.907, 2.671) | 2.191 (1.890, 2.401) | 1.628 (1.366, 1.903) | 3.010 (2.690, 3.339) | 2.402 (2.176, 2.633) | 2.597 (2.387, 2.804) |
| **Konica Minolta L*** | -1.818 (-2.103, -1.506) | -1.255 (-1.618, -0.958) | -1.746 (-2.093, -1.495) | 2.234 (1.892, 2.625) | 2.337 (2.019, 2.682) | 1.733 (1.468, 1.954) | 2.880 (2.594, 3.125) | 2.653 (2.407, 2.891) | 2.460 (2.264, 2.680) |

#### Supplemental Table 6. Variance per tertile in each skin tone measurement, after adjusting for covariates.

Standard deviation within each tertile of skin tone. In linear mixed-effects models on measurement bias of SpO_2_, where we use heterogeneous variation within tertiles(Supplemental Formula 9). Results indicate that within-group variation in the darkest skin group is consistently lower, after adjusting for covariates. Likelihood ratio tests indicate the performance of heterogenous variance models are consistently better across eight skin tone scales.

|  | **Standard deviation within tertile** | | |  | **LRT of full model vs reduced model** | |
| --- | --- | --- | --- | --- | --- | --- |
| **Skin Tone Variable** | **Lighter** | **Medium** | **Darker** | **N** | **Chi-Square** | **P-value** |
| Fitzpatrick | 2.24 | 1.90 | 1.64 | 463 | 11.63 | 0.003 |
| Von Luschan | 2.24 | 1.93 | 1.75 | 463 | 8.98 | 0.01 |
| Monk | 2.29 | 1.83 | 1.69 | 463 | 13.98 | 0.001 |
| Delfin ITA | 2.25 | 2.05 | 1.58 | 463 | 16.42 | <0.001 |
| Delfin Melanin Index | 2.09 | 2.18 | 1.57 | 463 | 15.37 | <0.001 |
| Delfin L* | 2.25 | 2.03 | 1.58 | 463 | 17.07 | <0.001 |
| Konica Minolta L* | 2.20 | 2.08 | 1.58 | 424 | 13.67 | <0.001 |
| Variable L* | 2.14 | 2.10 | 1.49 | 367 | 14.84 | <0.001 |

### Supplemental Figures

#### Supplemental Figure 1. Pairwise comparison of each skin tone scale distribution

Each skin tone variable is compared against each other, after normalization, with 2D hexagonal binning plots. Each plot was set to contain 10 full hexagons on the x-axis. A common legend is displayed in the right bottom corner, representing the color code of the hexagonal bin counts, across all subplots. As a sanity check, in the diagonal plots each skin tone variable is plotted against itself, where an identity relation is observed.

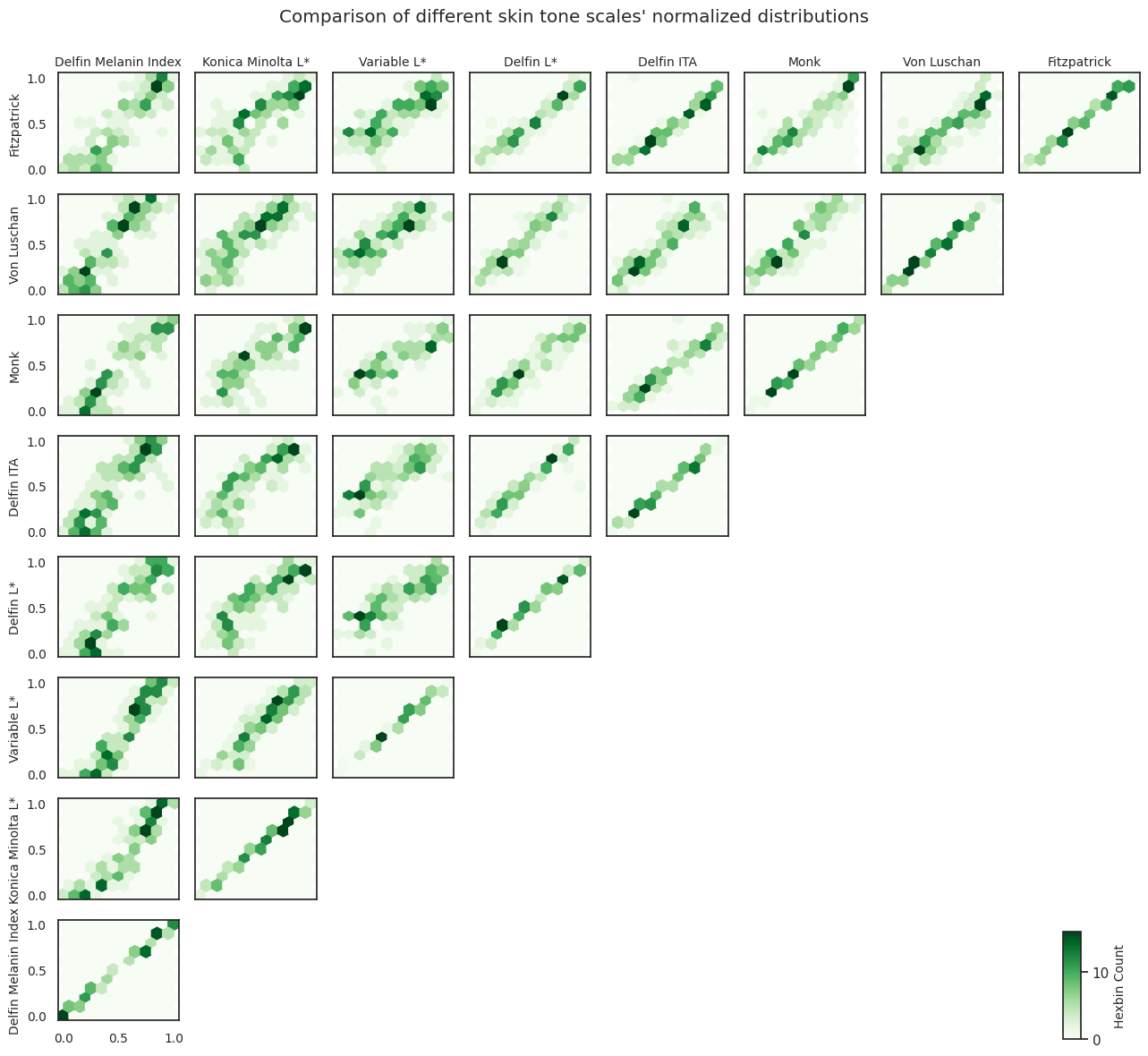

#### Supplemental Figure 2. Normalized Distribution of each considered skin tone scale, across all patients (N = 128)

The overlaid density plot represents skin tone distribution across the race groups. The X-axis is the relative proportion with 0 being the lightest observation and 1 being the darkest observation. The Y-axis represents the kernel density plots for each skin tone measurement.

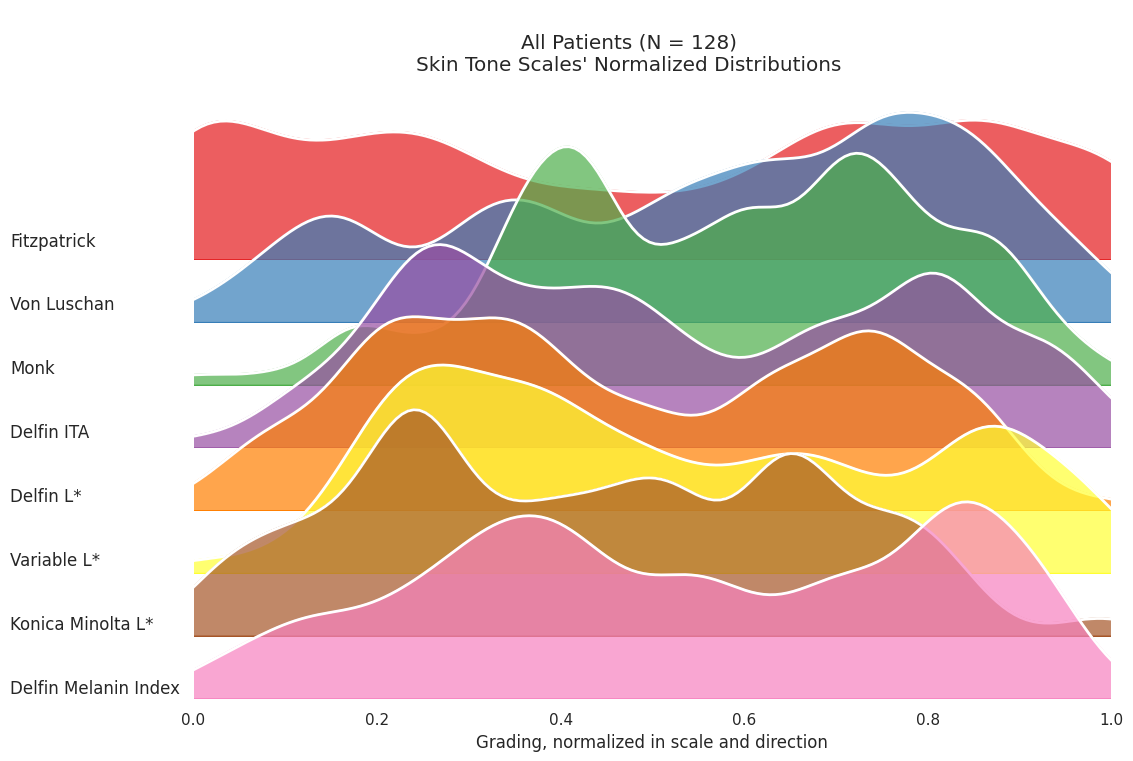

#### Supplemental Figure 3. Normalized Distribution of each considered skin tone.

The overlaid density plot represents skin tone distribution for the race group “Black” and “White”. The X-axis is the relative proportion with 0 being the lightest observation and 1 being the darkest observation.

##### Supplemental Figure 3a, among Black patients (N = 57))

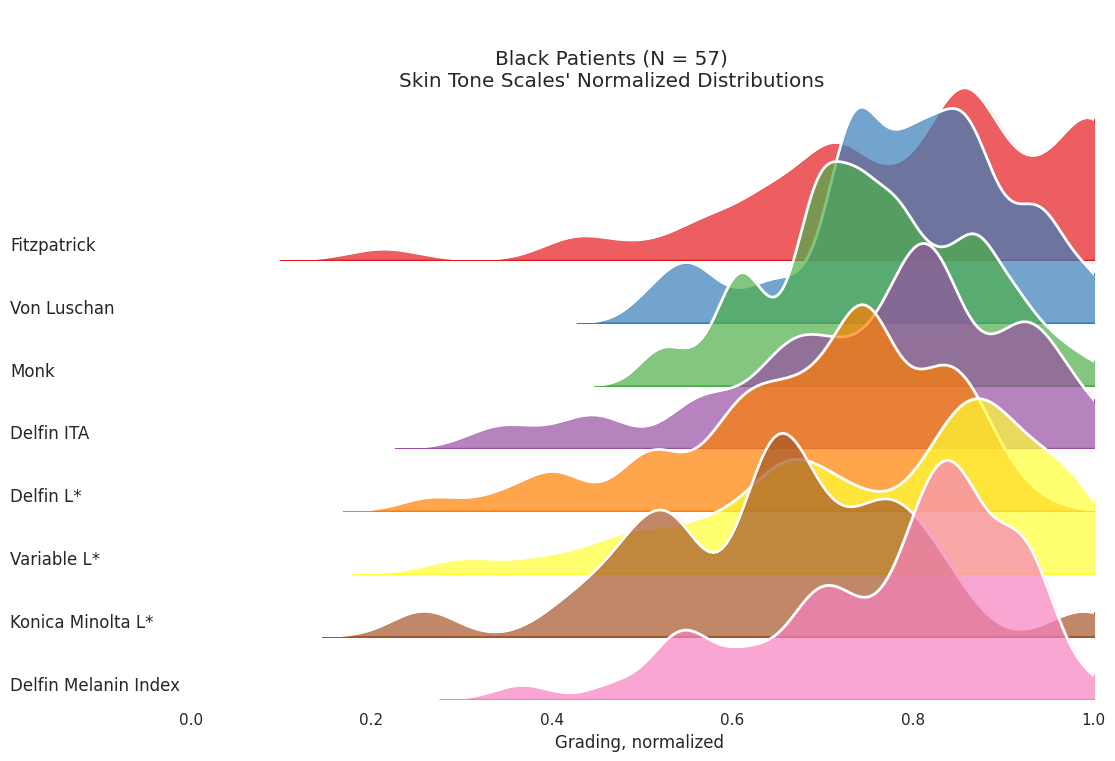

Supplemental Figure 3b. among White patients (N = 56)
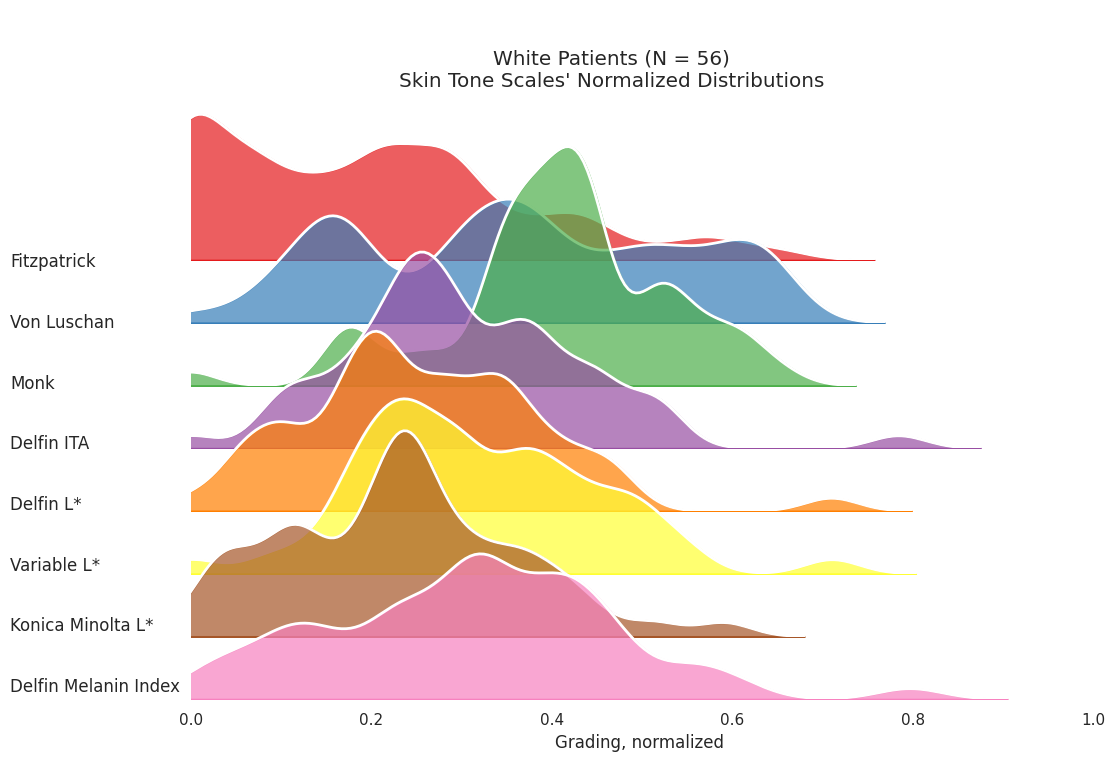

#### Supplemental Figure 4. Tertiles per skin tone covariate and race mapping

Distributions of skin tone data by race by tertiles are indicated in Supplemental Figure 3. Given a limited number of patients not of Black or White race, these patients are aggregated into “Other”. As expected, the majority of Black and White patients are of opposite tertiles. However, 9-19% are included in the median tertile. For White patients, 0.0-7.6% were included in the darkest tertile. For Black patients, 0.8-7.8% were in the lightest tertile.

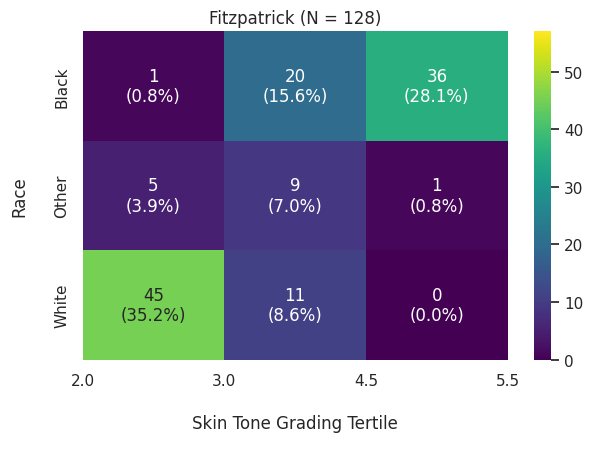

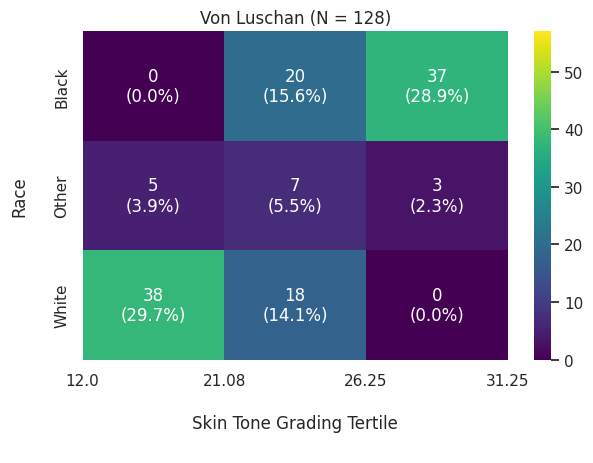

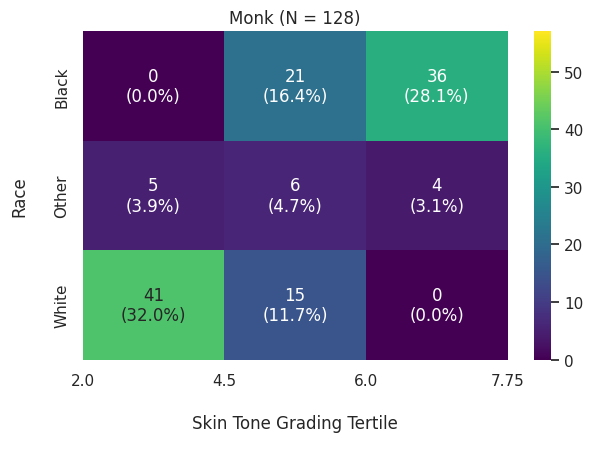

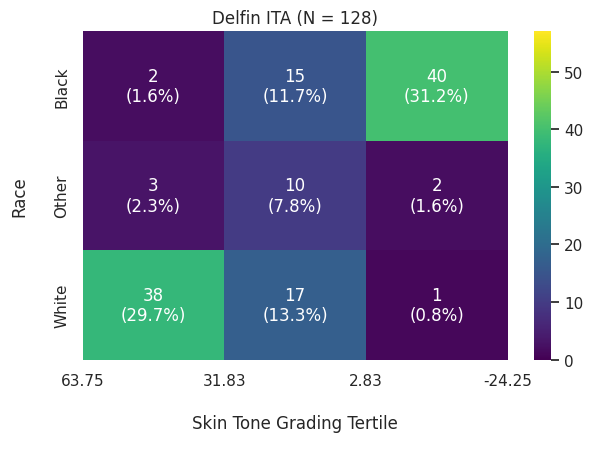

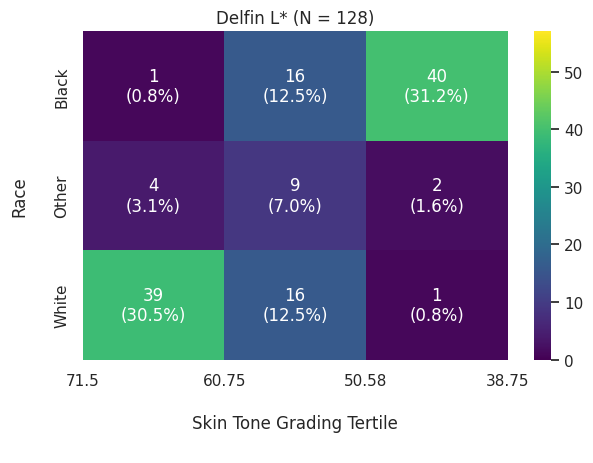

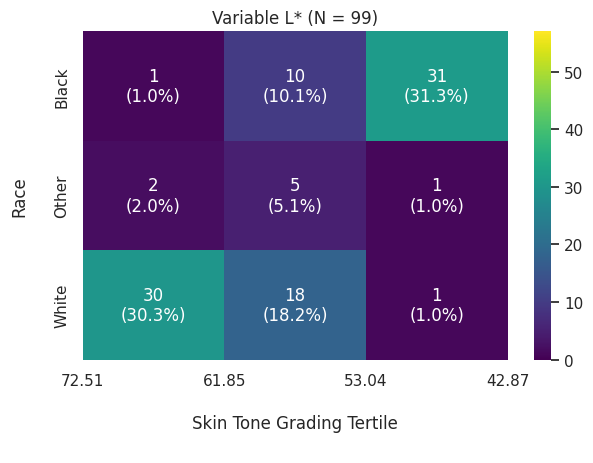

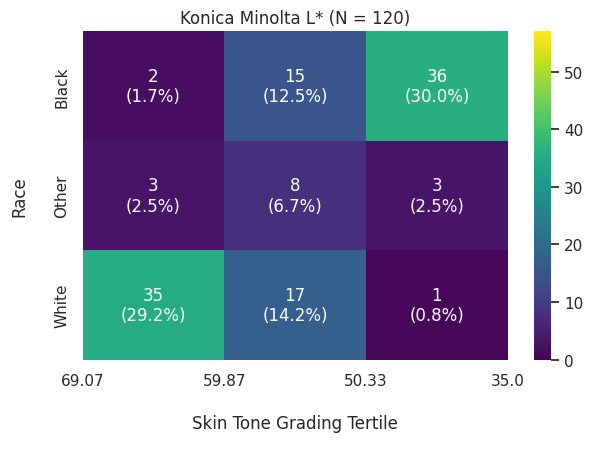

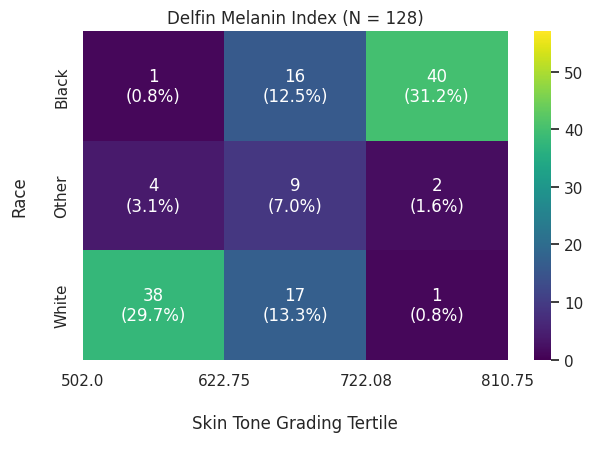

#### Supplemental Figure 5. Measured bias per SpO_2_ range and skin tone tertile

Boxplots of mean directional bias per SpO_2_, split by tertiles of skin tone. Each plot represents one skin tone scale, used as an axis of disparity.

Across all skin tone scales and SpO_2_ ranges, darker-pigmented patients tend to present a lower mean directional bias. In the lowest SpO_2_ range, lighter patients seem to undergo SaO_2_ underestimation, with a potential protective effect, which does not happen for darker patients. In the highest SpO_2_ range, the mean directional bias is negative and similar across skin tones. As we go from lower to higher SpO_2_ ranges, there is a trend of diminishing mean directional bias (from positive to negative) across all skin tones.

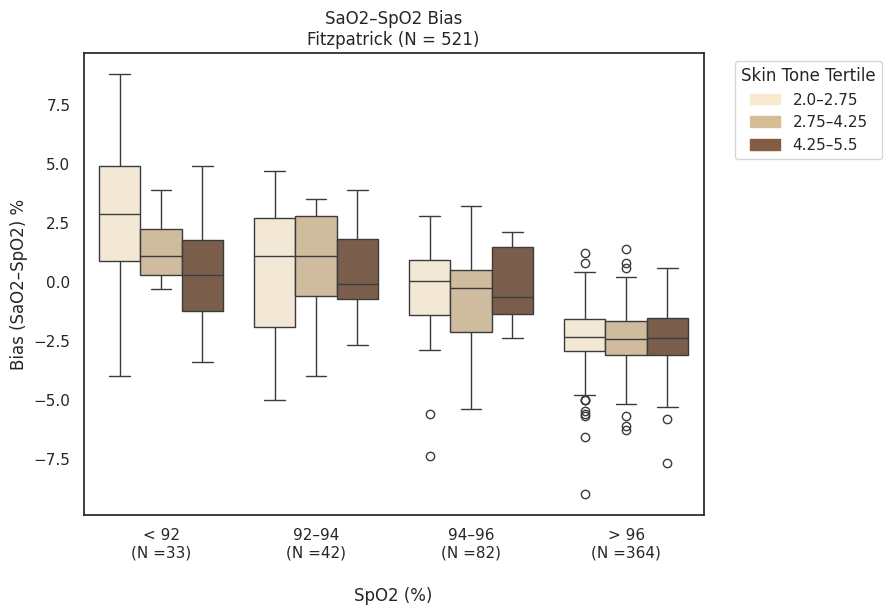

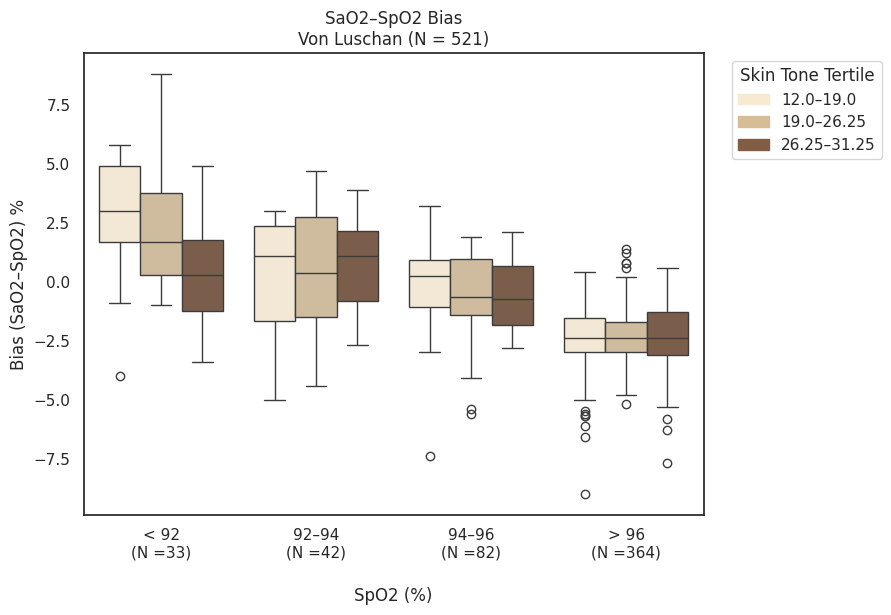

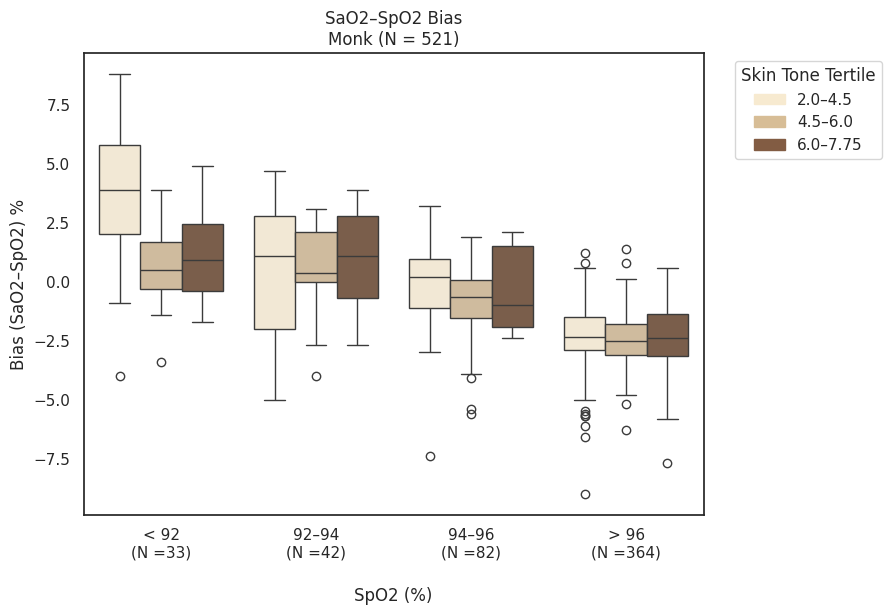

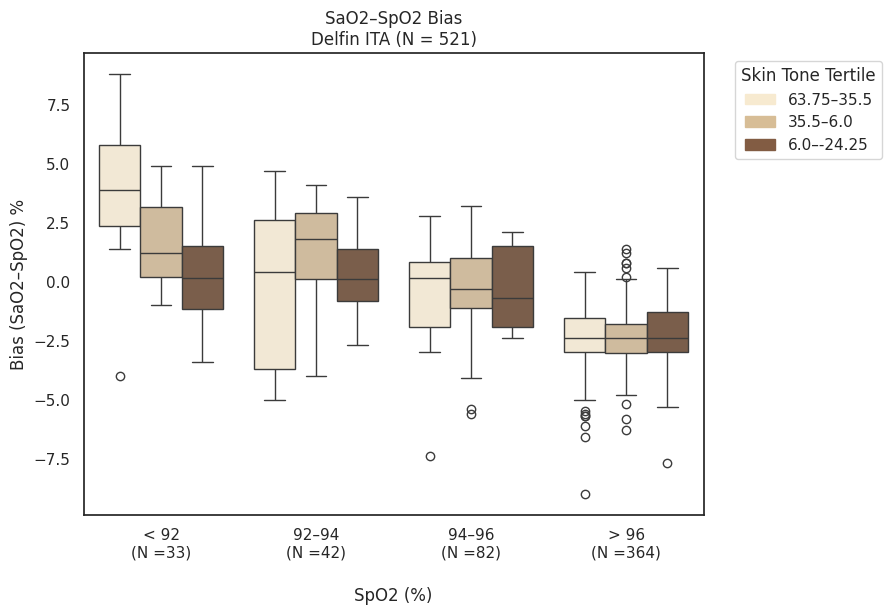

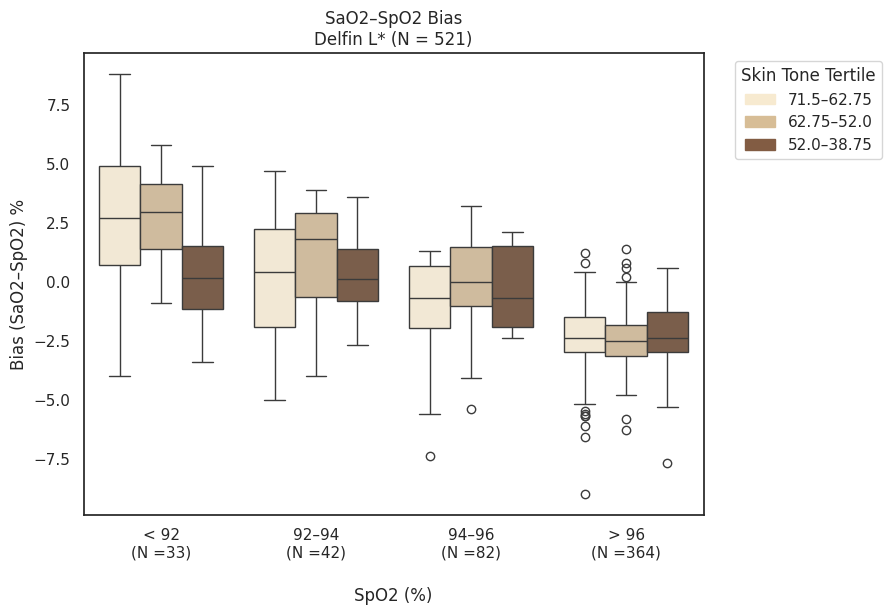

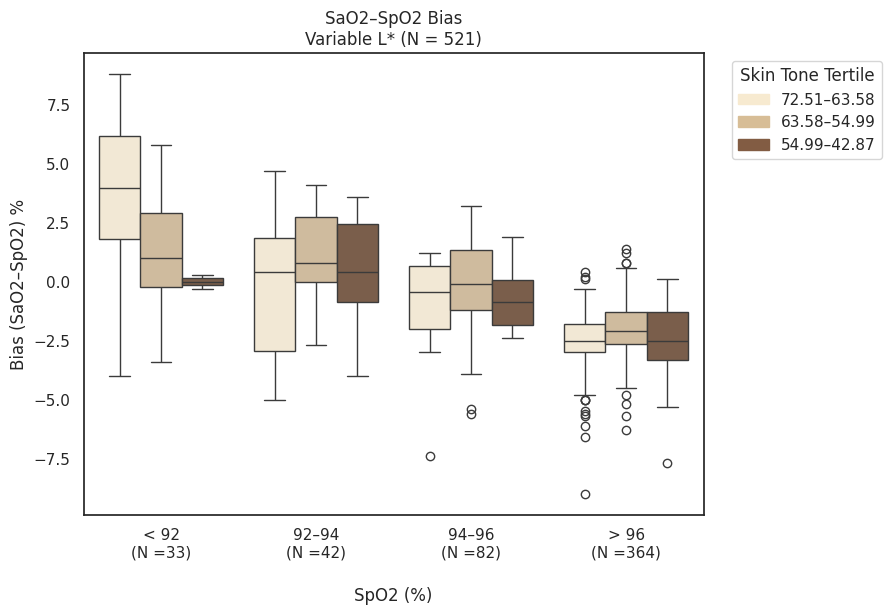

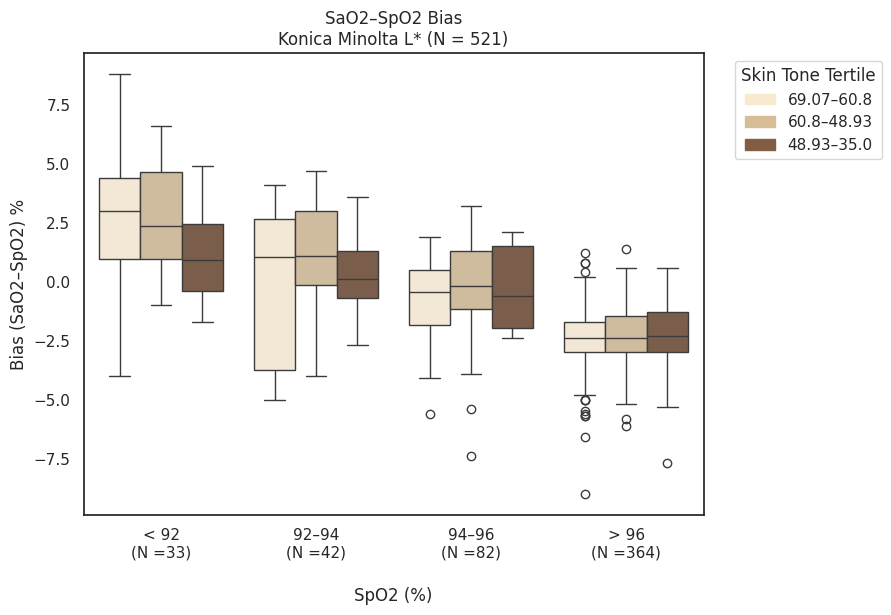

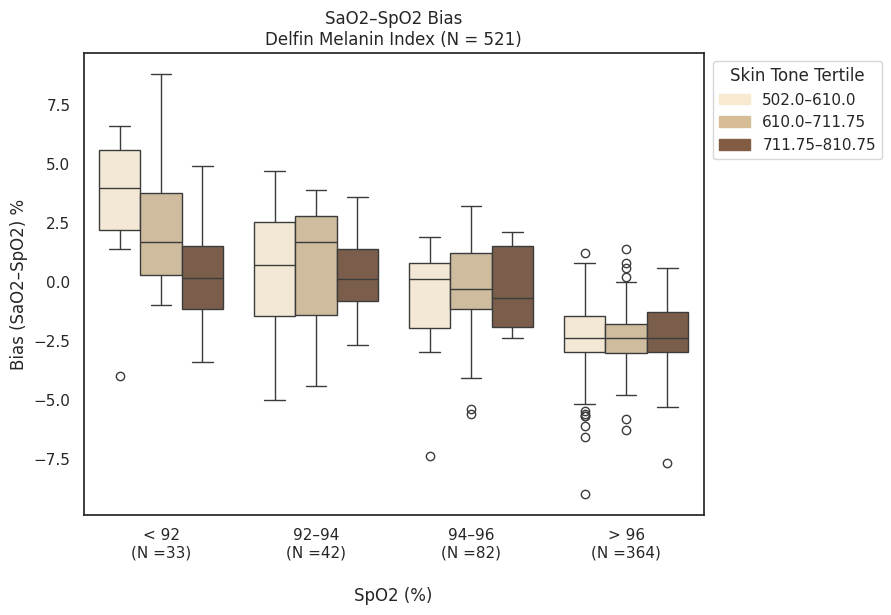

#### Supplemental Figure 6. Administered Skin Scales

##### Supplemental Figure 6a. Fitzpatrick Skin Scale.

Translated to color scales as per “Correlation between light absorbance and skin tone using fabricated skin phantoms with different colors” ^42^

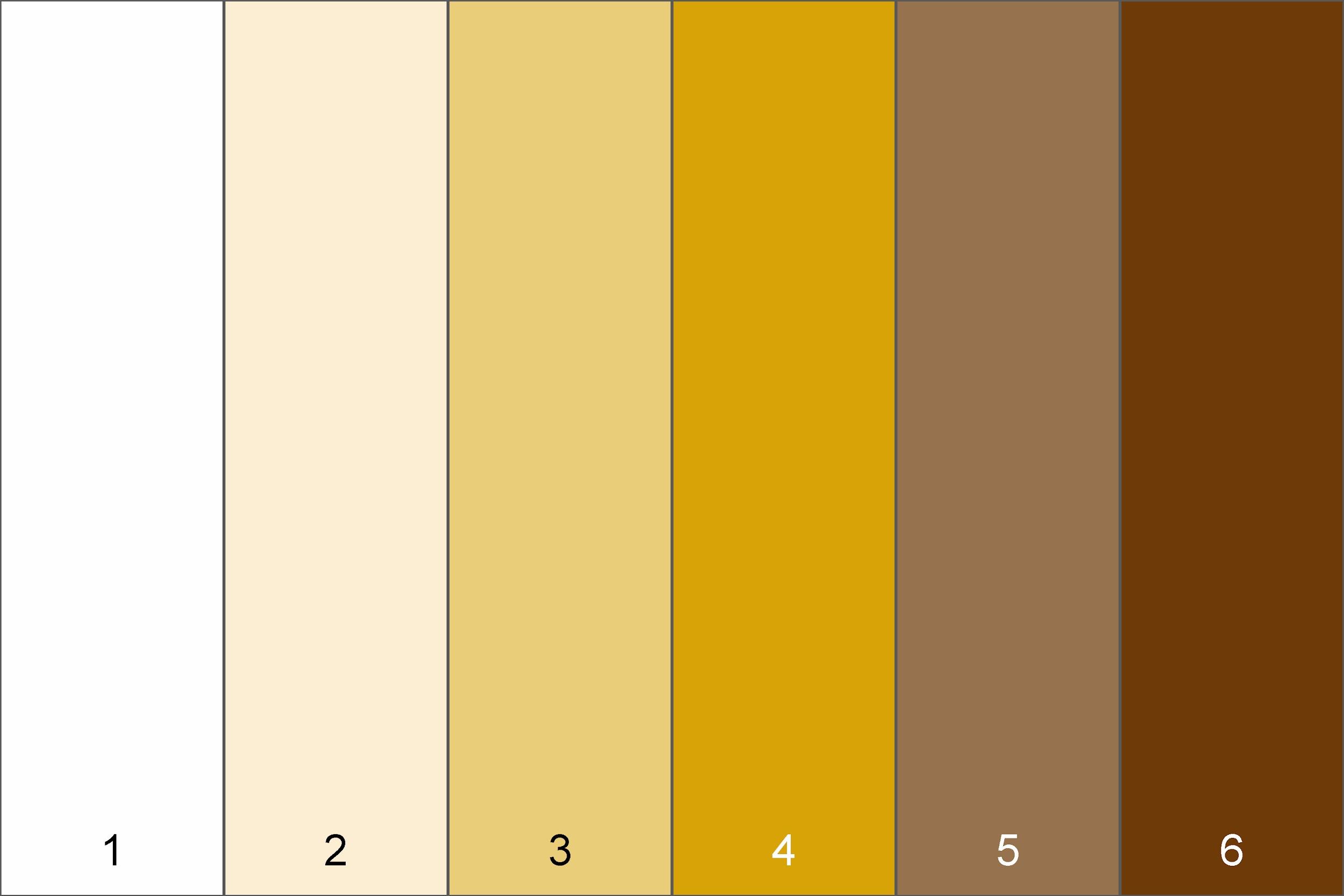

##### Supplemental Figure 6b. Von Luschan Chromatic Scale

From ^43^, reportedly replicated

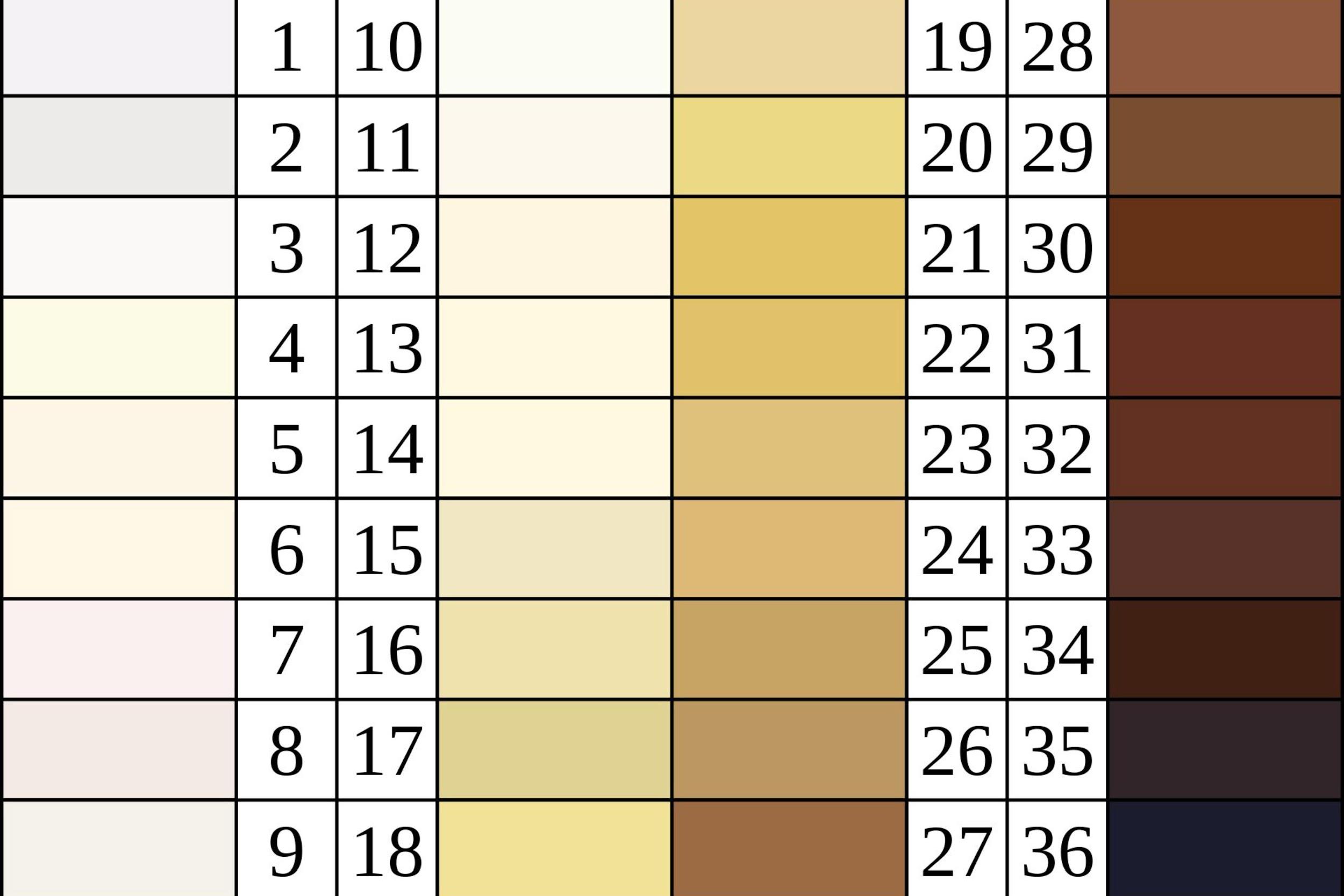

##### Supplemental Figure 6c. Monk Skin Tone Scale

Replicated from ^44^

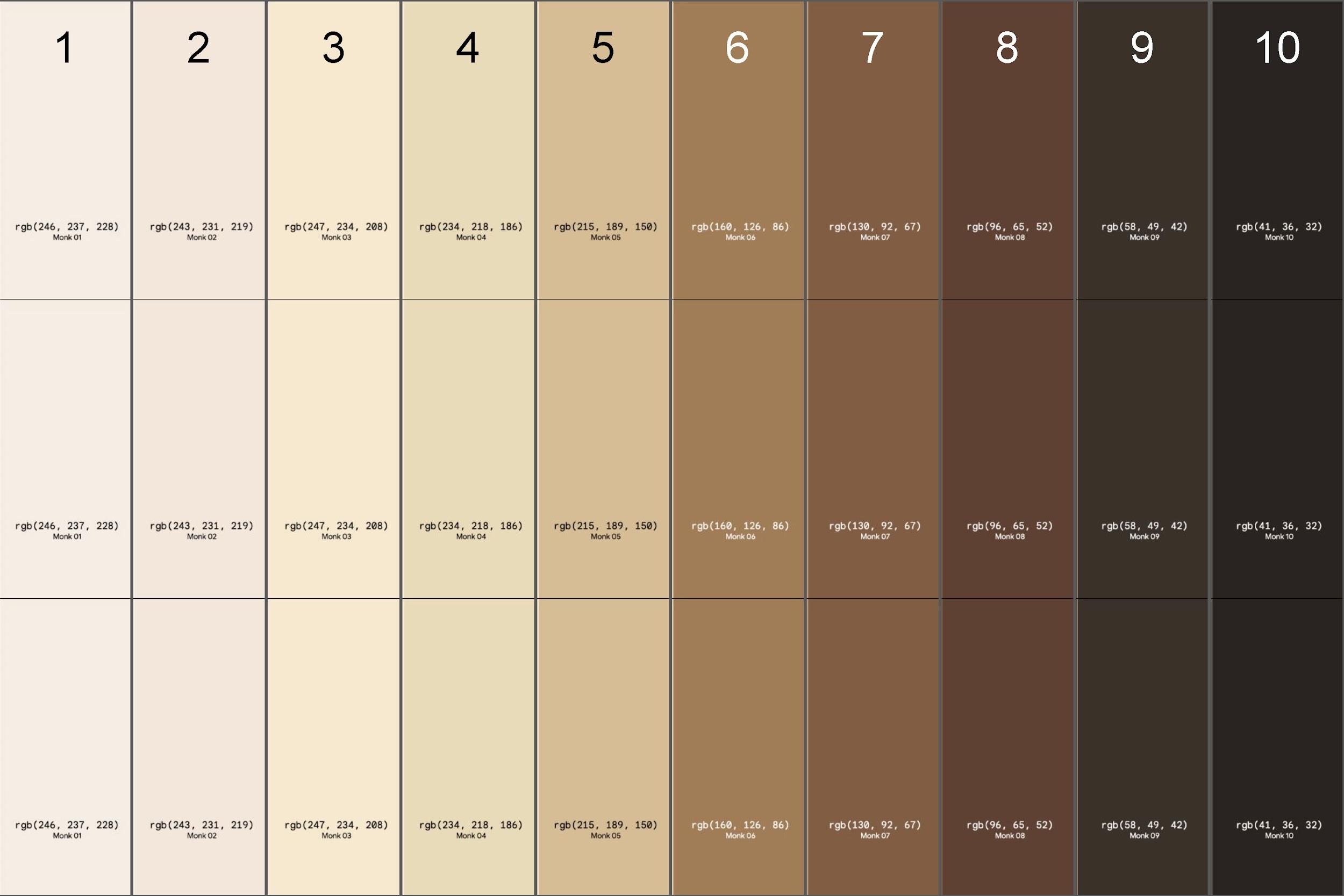

#### Supplemental Figures 7. Sensitivity Analyses of Figure 2.

##### Supplemental Figure 7a. Sensitivity Analysis | SaO₂ – SpO₂ | ≤ 6

Unadjusted error metrics of mean directional bias, standard deviation, and accuracy root mean square (also known as A_RMS_ or root mean square error), across tertiles. Tertiles are ordered from lightest to darkest, from the left to the right on the x-axis. Note that a pulse oximetry bias defined as SaO_2_ - SpO_2_ results in a negative bias reflecting that pulse oximetry overestimates true oxygenation values. Fitzpatrick and Monk appear to have a trend towards more negative bias (e.g., bias increasingly negative) from lighter to darker tertiles. A_RMS_ appears to be lower (that is, a lower root mean square error) in many darker tertiles than in lighter tertiles. Variable L* and Konica Minolta L* have fewer patients because there was more missingness (e.g., some patients did not have these measurements either due to patient refusal or interruptions by clinical workflow). The measurement of SpO_2_ is subject to variation. This sensitivity analysis was performed to eliminate the tertile trend due to outliers, with absolute values of SpO2 / SaO2 pairs larger than 6% being removed.

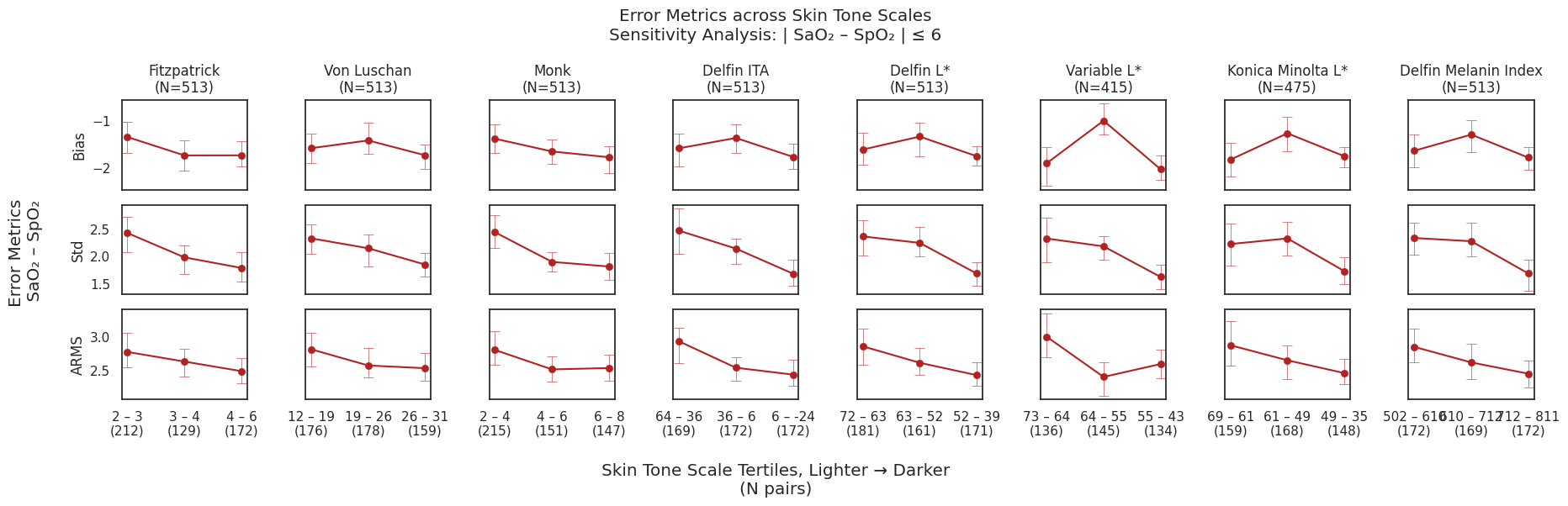

##### Supplemental Figure 7b. Sensitivity Analysis | SaO₂ – SpO₂ | ≤ 3.3 (= 1.5 IQR)

Unadjusted error metrics of mean directional bias, standard deviation, and accuracy root mean square (also known as A_RMS_ or root mean square error), across tertiles. Tertiles are ordered from lightest to darkest, from the left to the right on the x-axis. Note that a pulse oximetry bias defined as SaO_2_ - SpO_2_ results in a negative bias reflecting that pulse oximetry overestimates true oxygenation values. Fitzpatrick and Monk appear to have a trend towards more negative bias (e.g., bias increasingly negative) from lighter to darker tertiles. A_RMS_ appears to be lower (that is, a lower root mean square error) in many darker tertiles than in lighter tertiles. Variable L* and Konica Minolta L* have fewer patients because there was more missingness (e.g., some patients did not have these measurements either due to patient refusal or interruptions by clinical workflow). The measurement of SpO2 is subject to variation. This sensitivity analysis was performed to eliminate the tertile trend due to outliers, with absolute values of SpO2 / SaO2 pairs larger than 1.5 IQR removed.

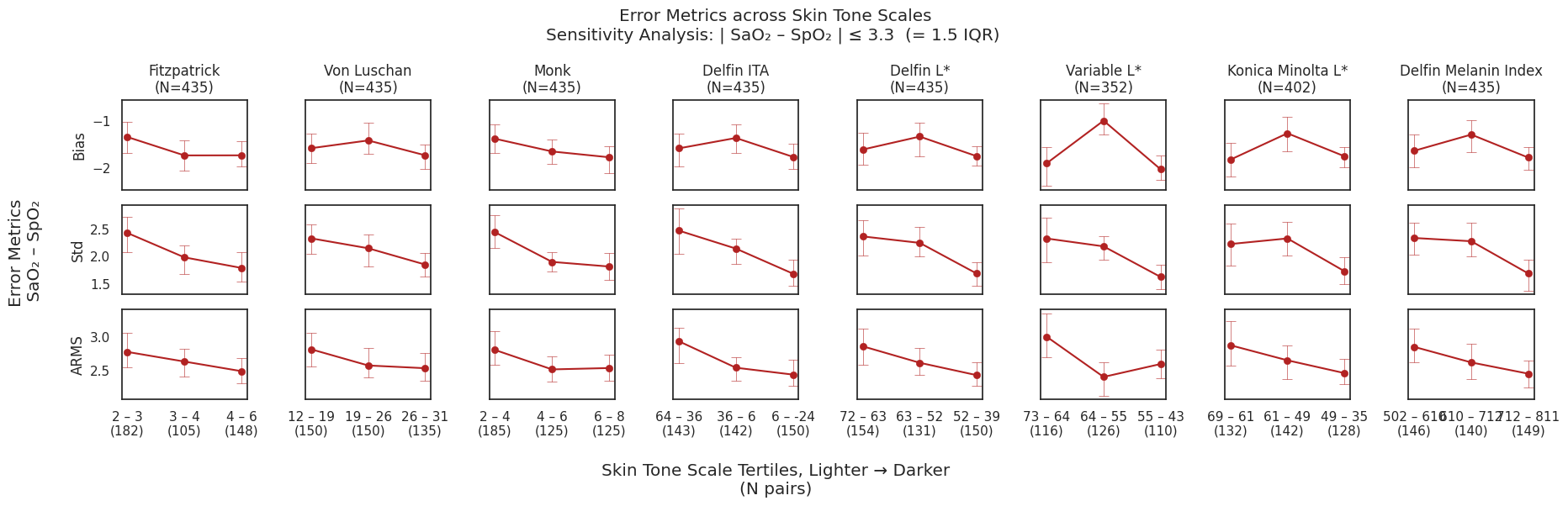
